# Activin A-activated ALK4 induces pathogenic Th17 involvement in endothelial–mesenchymal transition in systemic lupus erythematosus-associated pulmonary arterial hypertension

**DOI:** 10.1101/2025.01.23.25321014

**Authors:** Shuliang Jing, Junyan Qian, Mingxin Yao, Pei Mao, Jingyuan Zhang, Zhihong Wu, Hongjie Ying, Lie Wang, Mengtao Li, Jun Yang

**Affiliations:** Department of Physiology, Department of Cardiology of the Second Affiliated Hospital and School of Basic Medical Sciences, Zhejiang University School of Medicine, Hangzhou, 310058, China; Department of Rheumatology, Peking Union Medical College Hospital, Beijing, 100730, China; Stem Cell Facility of National Infrastructures for Translational Medicine, Institute of Clinical Medicine, Peking Union Medical College Hospital, Chinese Academy of Medical Sciences and Peking Union Medical College, Beijing, 100730, China; Zhejiang University School of Medicine, School of Basic Medical Sciences, Department of Immunology, Hangzhou, 310058, China; State Key Laboratory of Transvascular Implantation Devices, Hangzhou, 310009, China

## Abstract

**Objective:** Autoimmune diseases, such as systemic lupus erythematosus (SLE), are associated with pulmonary arterial hypertension (PAH), a condition that can lead to heart failure. However, whether T cells also contribute to the occurrence of PAH in SLE, has not been clarified. The objective of this study was to elucidate the role of Activin A signaling in the effector cells mainly involved in SLE-PAH.

**Methods:** CyTOF analysis was performed to identify the major affected immune cell population after the treatment in SLE-PAH patients. ELISA showed the serum Activin A and IL-17 levels were significantly higher in patients with SLE-PAH compared to SLE alone and healthy donors. We also conducted Th17 cells coculturing with pulmonary microvascular endothelial cells(PMECs) and constructed a SLE-PH mouse model and CD4^+^ T cells depletion mice together with two rat models to identify the converged target.

**Results:** The reduced CD4^+^ T cell number was detected in SLE-PAH patients after treatment. Activin A signals via ALK4 in both Th17 cells and PMECs. When ALK4 was overexpressed in Th17 cells, IL-6 and CTGF gene expression was significantly increased in cocultured PMECs. We found severe SLE-PH in mice by overexpression ALK4, and alleviated hemodynamic changes in CD4^+^ T cells depletion mice. ALK4 inhibitor TEW is effective to treat PAH by repressing CTGF transcription, which was facilitated by synergistic increases in pSmad2 and pSTAT3 levels downstream of ALK4 activation.

**Conclusion:** Our findings suggest that Activin A activates ALK4 in Th17 cells to induce IL-17 secretion, meanwhile activated ALK4 via Smad2 phosphorylation to induce EndoMT in hPMECs, indicating that ALK4 is a promising therapeutic target for SLE-PAH.

## Introduction

Autoimmune diseases, including connective tissue disease (CTD), are strongly associated with the development of irreversible end-stage pulmonary arterial remodeling, leading to pulmonary arterial hypertension (PAH; defined as a mean pulmonary artery pressure (mPAP) >20 mmHg) at rest[1]. Although PAH is related primarily to systemic sclerosis in Caucasian populations, systemic lupus erythematosus (SLE)-associated PAH is a more common form of CTD-PAH in Asian individuals[1]. Current treatments have yet to achieve complete cure for SLE-PAH, and the prognosis for the patients continues to be challenged [2, 3]. Therefore, identifying molecular targets closely related to the progression of SLE-PAH is important for the development of interventions.

In patients with SLE-PAH, the protein level of IL-17 in the lung and serum were found to be significantly greater than those in patients with SLE without PAH, and similar differences were observed between disease model and control mice[4, 5]. Th17 cells are the major source of IL-17; they are a subtype of CD4^+^ T cells associated with many inflammatory and autoimmune diseases, and they secrete IL-17 upon induction by cytokines such as TGF-β and IL-6, which contribute to the activation and differentiation of immune cells[6–8]. Furthermore, evidence indicates that upon exposure to cytokines such as IL-6, IL-1β and IL-23, Activin A is upregulated in CD4^+^ T cells, which induces their differentiation into pathogenic Th17 cells through IL-17 and GM-CSF production; these Th17 cells were found to exacerbate local inflammation in an animal model of autoimmune encephalomyelitis[7]. However, further investigation is necessary to determine whether the induction of pathogenic Th17 cell differentiation is involved in SLE-PAH.

The components of the TGF-β pathway, including bone morphogenetic protein type II receptor (BMPRII) and Activin Receptor-Like Kinase type 1(ALK1) have been implicated in idiopathic and heritable PAH[9, 10]. BMPRII plays a vital role in maintaining pulmonary vascular endothelial integrity, and mutations in its encoding gene have been detected in individuals with heritable PAH[11–13]. In contrast, Activin A, another TGF-β family member, and its receptor ALK5 coordinately mediate the abnormal proliferation of pulmonary vascular endothelial cells, which is related to elevated pulmonary artery pressure[14–17]. Patients with PAH have increased Activin A levels in peripheral blood and elevated levels of proinflammatory cytokines such as IL-6[18–20]. In this study, we investigated the onset of PAH in the context of SLE, especially the initiation of microvascular endothelial cell dysfunction upon specific immune insults.

Current treatments for SLE-PAH include endothelin receptor antagonists, phosphodiesterase type 5 inhibitors and prostacyclin analogs, which are often administered in combination. In 2023, a fourth potential therapy for PAH[1], sotatercept, which is a recombinant human ACTRIIA fusion protein that acts as a ligand trap for Activin A, was reported to significantly decrease pulmonary vascular resistance in PAH patients and improve their performance on a 6-minute walk test[21, 22]. However, whether this approach is effective for a broader population of CTD patients, especially those with SLE-PAH, which has a high regional incidence rate, is still unclear. Moreover, some adverse events, including bleeding events and telangiectasia, occurred in the phase III clinical trial referenced above, indicating that safer and specific therapeutic strategies for CTD-PAH must be considered.

In this study, we aimed to delineate the mechanism of Activin A-activated signaling in the inflamed pulmonary vasculature and to investigate the role of specific receptors in the pathogenic effects of Th17 cells involved in SLE-PAH. Our study revealed a significant reduction in the CD4^+^ T cells population in the SLE-PAH post-treatment group compared with the pre-treatment group and confirmed the involvement of CD4^+^ T cells in PH mice. The increased levels of IL-17 and Activin A in the peripheral blood and lung tissue of SLE-PAH patients promoted the activation of target genes such as CTGF in human pulmonary microvascular endothelial cells (hPMECs). We induced more severe pulmonary hypertension by overexpressing the Activin A receptor ALK4 in pristane-induced mice exposed to hypoxia. We treated the mice with a novel ALK4 inhibitor, TEW, and found a significant improvement in the hemodynamic index and vascular remodeling.

## 2. Methods and Materials

### 2.1 Patient enrollment

Before the study began, all individuals provided written informed permission by the principles established in the declaration of Helsinki approved by the Ethics Committee of Zhejiang University Medical College (No.014-2015). Patients with systemic lupus erythematosus-associated pulmonary arterial hypertension (SLE-PAH; total n = 45) and SLE patients without PAH (total n = 46, age- and sex-matched) was enrolled at Peking Union Medical College Hospital between November 2016 and May 2021. PAH was diagnosed based on the 2015 ESC/ERS guidelines by right-sided heart catheterization (RHC), with a mean pulmonary artery pressure (mPAP) of 25 mm Hg at rest and pulmonary vascular resistance (PVR) of >2 Wood units (WU)[23]. All patients with SLE were diagnosed using the American College of Rheumatology 1997 criteria[24]. Patients with additional overlapping CTDs, interstitial lung disease, pulmonary thromboembolism, left heart insufficiency, infection, or acute organ dysfunction were excluded.

### 2.2 Animal study design

All animal study protocols were in accordance with the ARRIVE 2.0 Guidelines[25]. All animal study protocols were reviewed and approved by Animal Care & Use Committee of Zhejiang University (Number: ZJU20210036 and ZJU20240927, AIRB-2023-0317). Animals were randomly assigned to the experimental groups. Following the principle of equal opportunity, the random number table method was used to assign each animal. No samples or animals were excluded from the analysis. All quantitative analyses and measurements (gene expression analysis, ELISA, RVSP measurement, WB analysis) were performed in a blinded manner. All figures are representative of at least three experiments unless otherwise noted. At the time of sacrifice, mice were anaesthetized with pentobarbital sodium (1% in 0.9% saline, 0.1 mL/10 g body weight) injected intraperitoneally once. Rats were anaesthetized with pentobarbital sodium (2% in 0.9% saline, 0.2 mL/100 g body weight) injected intraperitoneally once. The anaesthesia depth was monitored with pedal reflex. At the end of the experiment, all animals were euthanized by cervical dislocation.

### 2.3 SLE-PH mouse model establishment

Female C57 mice (18–20 g, Charles River, n = 8 mice per group) were randomly grouped according to the experiments. The mice in the SLE and SLE-PH groups were intraperitoneally injected with 0.5 mL of pristane (MCE, USA), while saline was also intraperitoneally administered to the mice in the normoxia and hypoxia groups. Then, the mice in the hypoxia and SLE-PH groups were placed in a 10% O2 chamber and maintained under hypoxic conditions for four weeks to establish the SLE-PH model.

All animals were housed in an SPF facility (3–4 mice per cage) in a room with a temperature of 25 ± 2°C, were maintained on a 12 h light/dark cycle, and were allowed free access to a standard rodent chow diet and water. At the endpoint of model establishment, the Vevo 2100 system (VisualSonics, Toronto, ON, Canada) was used for transthoracic echocardiography.

### 2.4 Statistical analysis

All the statistical analyses were performed using GraphPad Prism 8 (GraphPad Software, San Diego, USA). All the data were analyzed without the exclusion of any data points. All experimental data were subjected to a normality (Shapiro–Wilk) test. Unless otherwise noted, statistical comparisons of samples were conducted using Student’s t test or one-way repeated measures ANOVA with the Dunn–Bonferroni correction for multiple comparisons followed by Dunnett’s post hoc test. For nonnormally distributed data, statistical comparisons of samples were conducted using the Mann Whitney U test. P values < 0.05 (two-tailed) were considered to indicate statistical significance. In the corresponding figures, *, **, and *** indicate P<0.05, P<0.01 and P<0.001, respectively. Each figure legend mentions the test used, the criteria for statistical significance, and the number of experimental and biological replicates.

## 3. Results

### 3.1. The Activin A level was elevated in patients with SLE-PAH, which drove the expression of *IL-17A* and *GM-CSF* in CD4^+^ T cells

To identify the major affected immune cell type in SLE-PAH patients, we applied mass cytometry to determine the proportions of CD4^+^ T cells, CD8^+^ T cells, B cells and myeloid dendritic cells in the peripheral blood of patients with SLE-PAH before and after 6 months of treatment with a vasodilator or an immunosuppressant (Figure 1A; Supplemental Figure 1A). The treatment for these patients were listed in Supplemental Table 1. Compared to that in the pre-therapy group, the proportion of CD4^+^ T cells was significantly reduced in post-therapy SLE-PAH patients and was also reduced compared with the proportions of CD8^+^ T cells, B cells and myeloid dendritic cells, indicating that CD4^+^ T cells might be the major immune cells involved in the pathogenesis of SLE-PAH (Figure 1B and Supplemental Figure 1B).

**Figure 1.**
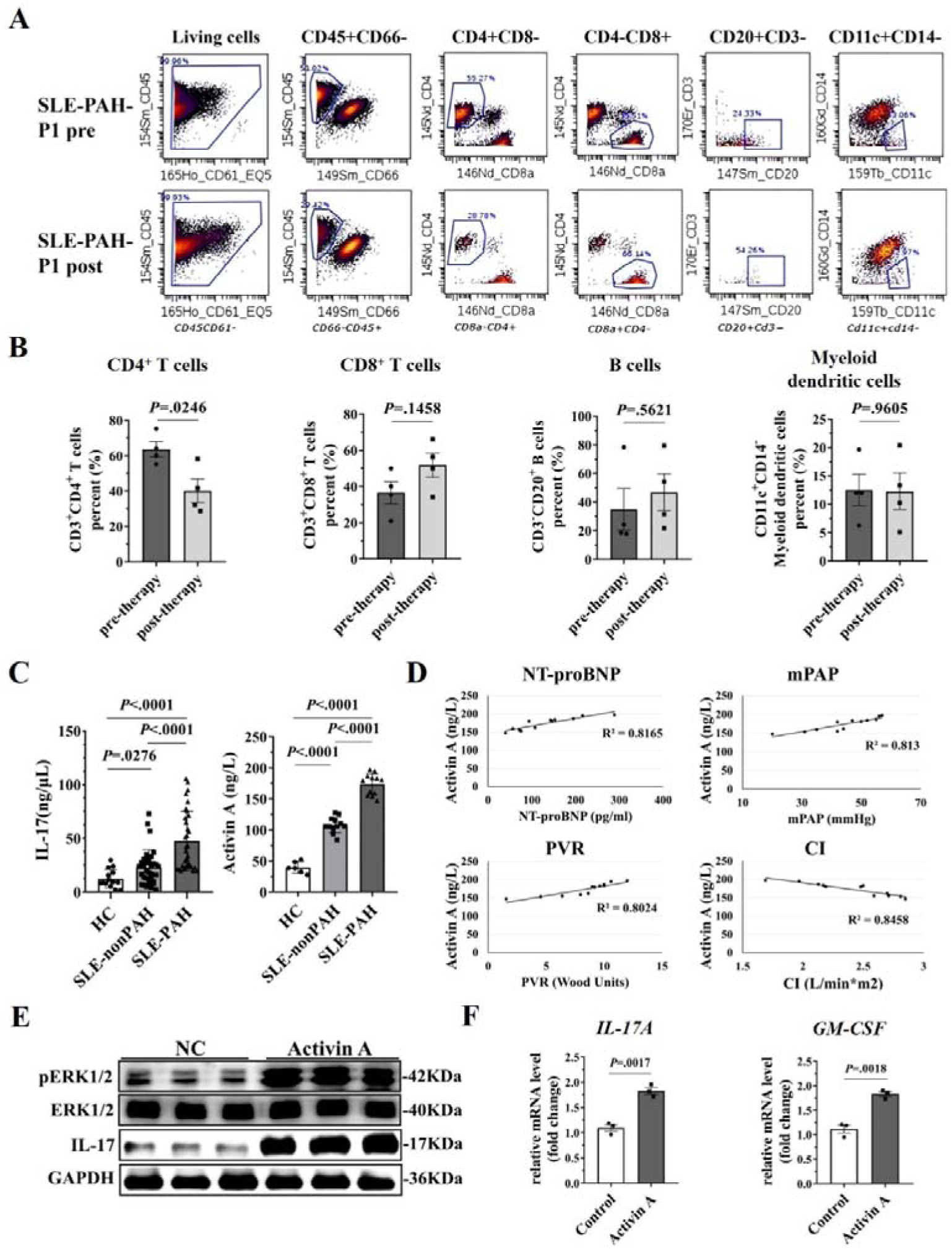
The Activin A level was elevated in patients with SLE-PAH, which drove the expression of *IL-17A* and *GM-CSF* in CD4^+^ T cells. **(A)** The percentages of CD4^+^ T cells, CD8^+^ T cells, B cells and Myeloid dendritic cells in the peripheral blood of the patients with pre- and post-therapy were analyzed by mass cytometry. **(B)** The quantification of analyzed cells from the four patients. **(C)** ELISA Assay were conducted with sera from healthy control, SLE-nonPAH patients (N=46) and SLE-PAH patients (N=45) “HC”: Healthy control. **(D)** Correlation analysis between Activin A level and clinical parameters in patients with SLE-PAH. Correlation was observed between Activin A and NT-proBNP, mPAP, PVR and CI. mPAP, mean pulmonary arterial pressure; PVR, pulmonary vascular resistance; CI, cardiac index. **(E)** The human PBMC-derived CD4^+^ T cells stimulated by Activin A (25ng/ml) for 18 h were collected and examined by western blot for p-ERK (left) and densitometric quantification was shown in right (N=3). **(F)** The mRNA expression of *IL-17A* and *GM-CSF* in human CD4^+^ T cells activated by Activin A (N=3). Data were shown as mean ±SEM, *P* values (Bonferroni corrected) are depicted in the panels throughout all figures as \**P* < .05, \*\**P* < .01 and \*\*\**P* < .001. ns, not significant. Using one-way ANOVA with repeated measures followed by Bonferroni correction for multiple comparisons.

To investigate the potential involvement of IL-17 and Activin A in SLE-PAH pathogenesis, we measured the levels of IL-17 and Activin A in serum from 33 healthy controls, 34 SLE- and 27 pSS-patients without PAH and 33 SLE-PAH and 24 pSS-PAH by ELISA. Our findings revealed that the IL-17 and Activin A levels were significantly higher in SLE-non and pSS-non PAH patients than in healthy controls and that Activin A levels were even higher in patients with SLE-PAH and pSS-PAH, indicating the potential role of Activin A in PAH progression in patients with CTD (Figure 1C and Supplemental Figure 1C). Similarly, as reported for other conditions associated with PAH, IL-17 levels were also higher in patients with CTD-PAH than in healthy controls.

Then we investigated the relationship between Activin A levels and key clinical parameters in patients with SLE-PAH. Our correlation analysis revealed a strong association between Activin A and NT-proBNP (R^2^= 0.8165), Activin A and mPAP (R^2^= 0.813), Activin A and PVR (R^2^= 0.8024), Activin A and CI (R^2^= 0.8458) (Figure 1D). We also found IL-17 had strong correlation with the above clinical parameters (Supplemental Figure 1D). These findings highlight the potential of Activin A and IL-17 as an important biomarker for assessing disease severity in SLE-PAH and underscore the need for further research to elucidate its role in the pathophysiology of this condition and its potential as a therapeutic target.

To examine the effect of enhanced Activin A on human Th17 cells differentiation, we isolated CD4^+^ T cells from human PBMCs and stimulated with Activin A for 18 h to investigate IL-17 expression and ERK phosphorylation, which is essential for pathogenic Th17 cell differentiation as previously reported[7]. We confirmed that CD4^+^ T cells differentiated into pathogenic Th17 cells with immunoblotting (Figure 1E and Supplemental Figure 1E). We also detected increased *IL-17A* and *GM-CSF* levels at mRNA level (Figure 1F).

These findings suggested that in the context of SLE-PAH, Activin A induced *IL-17* expression and release in CD4^+^ T cells.

### 3.2. The Activin A receptor ALK4 mediated the release of IL-17 from Th17 cells and induced EndoMT in cocultured hPMECs

Activin A binds to naïve CD4^+^ T cells to drive them to differentiate into pathogenic Th17 cells may through activation of the type I receptor ALK4. At the same time, activin A could also effect on endothelial cells. Considering the complexity of the inflammatory conditions in PAH, we investigated the interaction between PMECs and Th17 cells by coculture assays. In these assays, we further examined the role of the possible key player ALK4 by modulating its expression on Th17 cells to confirm its effect on cocultured PMECs[7]. We isolated CD4^+^ T cells from WT C57BL/6J mice, treated these cells with ALK4-shRNA, scramble-shRNA or ALK4-OE lentiviruses with or without Activin A in the presence of IL-6, and then cocultured the cells with PMECs isolated from WT mice (Figure 2A). The knockdown efficiency of ALK4-shRNA in Th17 cells was evaluated by qPCR and immunoblotting, as the efficiency of ALK4 overexpression (Figure 2B and Supplemental Figure 2). Further analysis of gene expression in Th17 cells revealed that the levels of *IL-17*, *GM-CSF* and *RORC* (a transcription factor for Th17 cells) were elevated in the ALK4-OE group[26] and that the level of the fourth marker of pathogenic Th17 cells, *IL-10*, was increased in the ALK4-shRNA group compared with the ALK4-OE and scramble-shRNA groups, respectively (Figure 2C).

**Figure 2.**
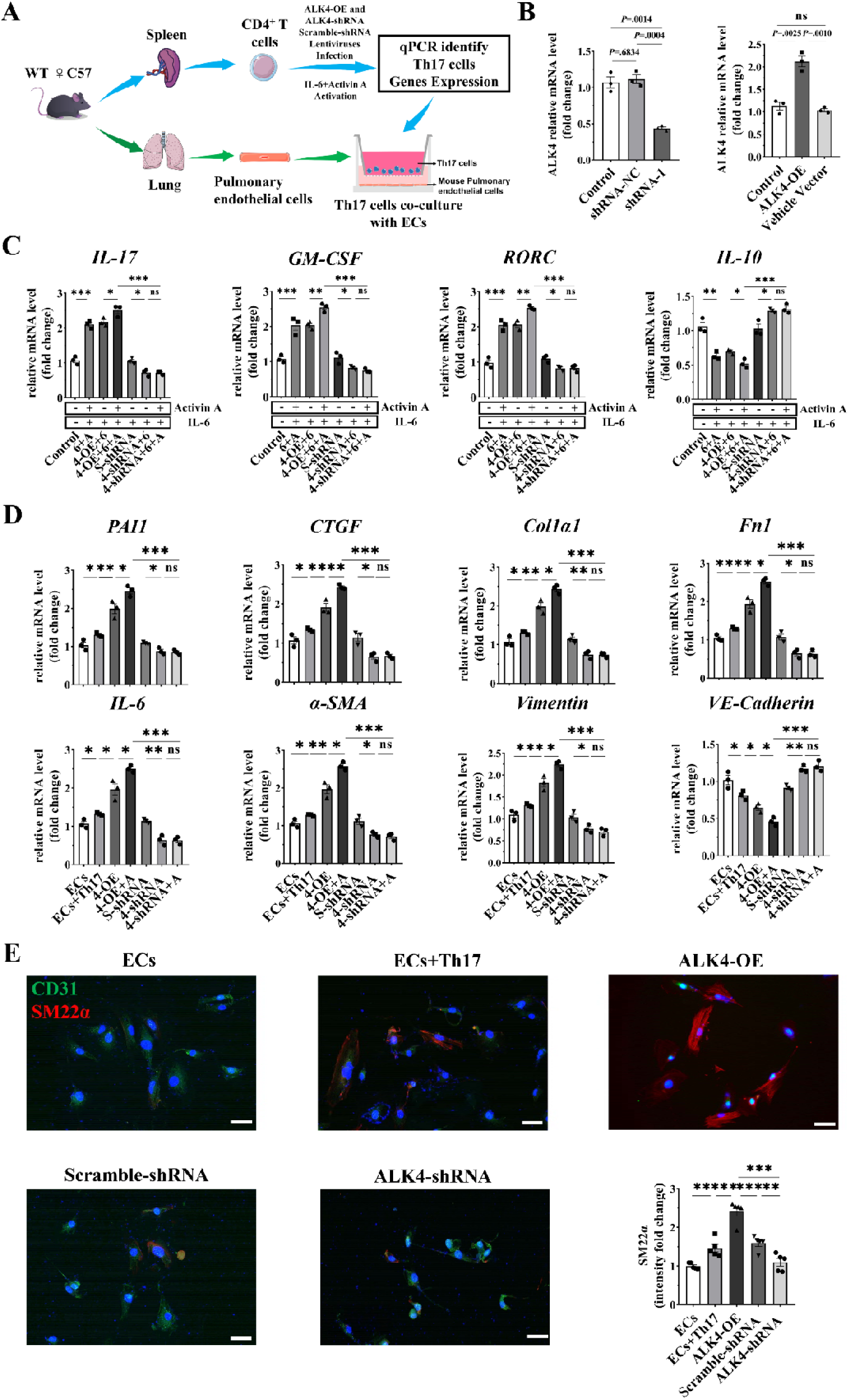
The Activin A receptor ALK4 mediated the release of IL-17 from Th17 cells and induced EndoMT in cocultured PMECs. **(A)** Schematic illustration of CD4+ T cells interaction with hPMECs. **(B)** The mRNA expression of *ALK4* after treated with ALK4-shRNA and ALK4-OE lentiviruses in CD4+ T cells. (N=3). **(C)** The mRNA expression in CD4+ T cells treated with ALK4-shRNA, ALK4-OE and scramble-shRNA lentivirus after 48 h (N=3). “6” indicated IL-6, “A” indicated Activin A. **(D)** The mRNA expression in mouse PMECs co-cultured with Th17 cells (N=3). **(E)** Representative images showed CD31 (green) and SM22α (red) with DAPI (blue) counterstain for nuclei in mouse pulmonary endothelial cells co-cultured with Th17 cells assessed by immunofluorescence microscopy from the indicated groups. Scale bar=50 μm. SM22α was quantified using ImageJ (right) (N=5). “ECs”: Endothelial cells. “OE”: overexpression. Data were shown as mean ±SEM, *P* values (Bonferroni corrected) are depicted in the panels throughout all figures as \**P* < .05, \*\**P* < .01 and \*\*\**P* < .001. ns, not significant. Using one-way ANOVA with repeated measures followed by Bonferroni correction for multiple comparisons.

To further confirm that the interaction between Th17 cells and PMECs is mediated by ALK4, we co-cultured PMECs and Th17 cells for a prolonged period. We found that the expression of genes related to Activin A-ALK4 signaling, such as *CTGF* and *PAI1,* ECM-related genes *Col1α1* and *Fn1*, inflammatory factor *IL-6* and mesenchymal markers, including α*-SMA* and *Vimentin* were significantly increased in the ALK4-OE co-culture group, but significantly decreased in the ALK4-shRNA co-culture group. Oppositely, the expression of *VE-Cadherin*, an endothelial marker, was decreased in the ALK4-OE co-culture group and increased in the ALK4-shRNA co-culture group (Figure 2D). We also examined smooth muscle cell marker SM22α expression in mouse pulmonary endothelial cells determined by immunofluorescent staining. We demonstrated that SM22α was highly expressed in ALK4-OE co-culture group and decreased in ALK4-shRNA co-culture group (Figure 2E).

These data suggest that Activin A exerts its effects through ALK4 to drive the differentiation of pathogenic Th17 cells, which release IL-17 to induce inflammation and detrimental EndoMT in PMECs.

### 3.3 CD4^+^ T cell depletion alleviated hemodynamic changes in hypoxia mice

To further investigate the involvement of CD4^+^ T cells in SLE-PAH, we generated CD4^+^ T cell-depleted mice by using anti-CD4 antibody GK1.5 for WT C57/BL6J mice and isotype antibody for control mice under chronic hypoxia (10% O_2_) exposure for 3 weeks (Figure 3A). The effectiveness of CD4^+^ T cells depletion was confirmed by a reduction in CD4^+^ T cells from approximately 13% to 5% of total splenic cells examined by flow cytometry (Figure 3B). Compared with the hypoxia group, CD4^+^ T cell-depleted mice exhibited a significant decrease in RVSP, TPVRI, and Fulton index (Figure 3C-E). The serum level of IL-17 was also reduced in CD4^+^ T cell-depleted mice (Figure 3F). However, CD4^+^ T cell depletion had less effect at reversing remodeled small pulmonary arteries with only partially reduced wall thickness (Figure 3G-I). A marked decrease in IL-17 levels was observed in the CD31+ pulmonary vasculature endothelium cells in CD4^+^ T cell-depleted mice (Figure 3I). Furthermore, immunoblotting with homogenized lungs demonstrated that a reduction in IL-6 and Smad2 phosphorylation, which was accompanied by decreased CTGF in CD4^+^ T cell-depleted mice (Figure 3J). The analysis of genes expression in lungs showed decreased expression levels of *IL-17A*, *GM-CSF*, *Rorc*, *CTGF*, *Col1α1*, *IL-6*, *Fn1* in CD4^+^ T cell-depleted mice (Supplemental Figure 3A).

**Figure 3.**
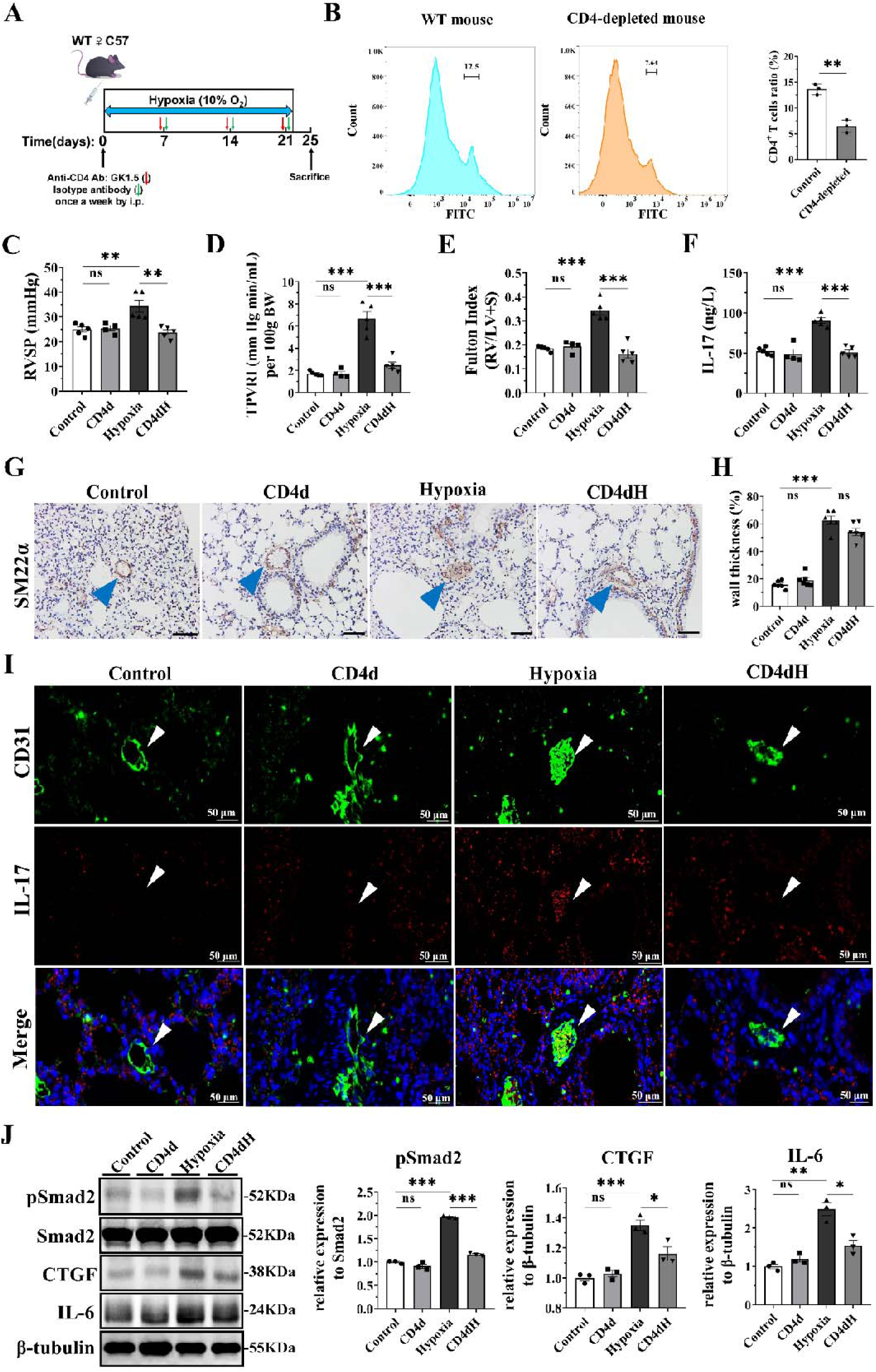
CD4-depletion alleviates hemodynamic changes in hypoxia mice. **(A)** The schematic diagram of constructing chronic hypoxia model in CD4-depleted mice. **(B)** CD4^+^ T cells depletion efficiency. Representative flow cytometry histogram of CD4^+^ T cells from spleens and quantification of CD4^+^ T cells (N =3). **(C, D, E)** Pulmonary hypertension parameters were as follows: RV systolic Pressure (RVSP), total pulmonary vascular resistance (TPVRI) and Fulton index was measured in the lungs of the SLE-PH mouse model. (N=6 mice for each group). **(F)** ELISA showing IL-17 level in the serum of different groups. **(G)** The representative images of pulmonary arteries (Scale bar=50 μm) by IHC staining of SM22α of each group indicated in the panel. Blue arrow indicates small pulmonary arteries **(H)** Medial wall thickness of small pulmonary arteries. **(I)** Representative images showed CD31 (green) and IL-17 (red) with DAPI (blue) counterstain for nuclei in SLE-PH mice. White arrow indicates the pulmonary small arterial. Scale bar=50 μm. **(J)** The western blot result showed that pSmad2, CTGF and IL-6 expression in lungs of each group (left) and densitometric quantification was shown in right (N=3). “CD4d”: CD4-deleted, “CD4dH”: CD4-depleted and hypoxia. The samples derive from the same experiment and that blots were processed in parallel. Data were shown as mean ±SEM, *P* values (Bonferroni corrected) are depicted in the panels throughout all figures as \**P* < .05, \*\**P* < .01 and \*\*\**P* < .001. ns, not significant. Using one-way ANOVA with repeated measures followed by Bonferroni correction for multiple comparisons.

These findings demonstrate that CD4 depletion confers protection against inflammation and hemodynamic change in hypoxia induced PH, whereas has less effect at reversal of vascular remodeling.

### 3.4. Aberrant activation of ALK4 but not ALK5 induced proinflammatory factor and EndoMT gene expression in hPMECs

Then to detect whether the downstream signaling pathway of Activin A is altered in hPMECs in the context of SLE-PAH, we examined the protein expression profiles of hPMECs stimulated with patient serum or Activin A (Figure 4A and Supplemental Figure 4A). We found that the phosphorylation of Smad2, a downstream mediator of the TGF-β pathway components, IL-6 and CTGF, which are known to induce EndoMT [28], were increased in hPMECs after incubation with SLE-non PAH patient serum, SLE-PAH patient serum or Activin A. Increased protein expression was induced by exposure to SLE-PAH patient serum compared with exposure to SLE-non PAH patient serum. To confirm Activin A dependent effect, we used Follistatin, a natural Activin-A antagonist, to block Activin A binding to its receptor[15, 27] and found reduced Smad2 phosphorylation, IL-6 and CTGF expression upon Activin A co-treatment with Follistatin comparing with no Follistatin. Therefore, the above results suggested that the Activin A/pSmad2 axis was activated in endothelial cells after stimulation with serum from patients with SLE-PAH.

**Figure 4.**
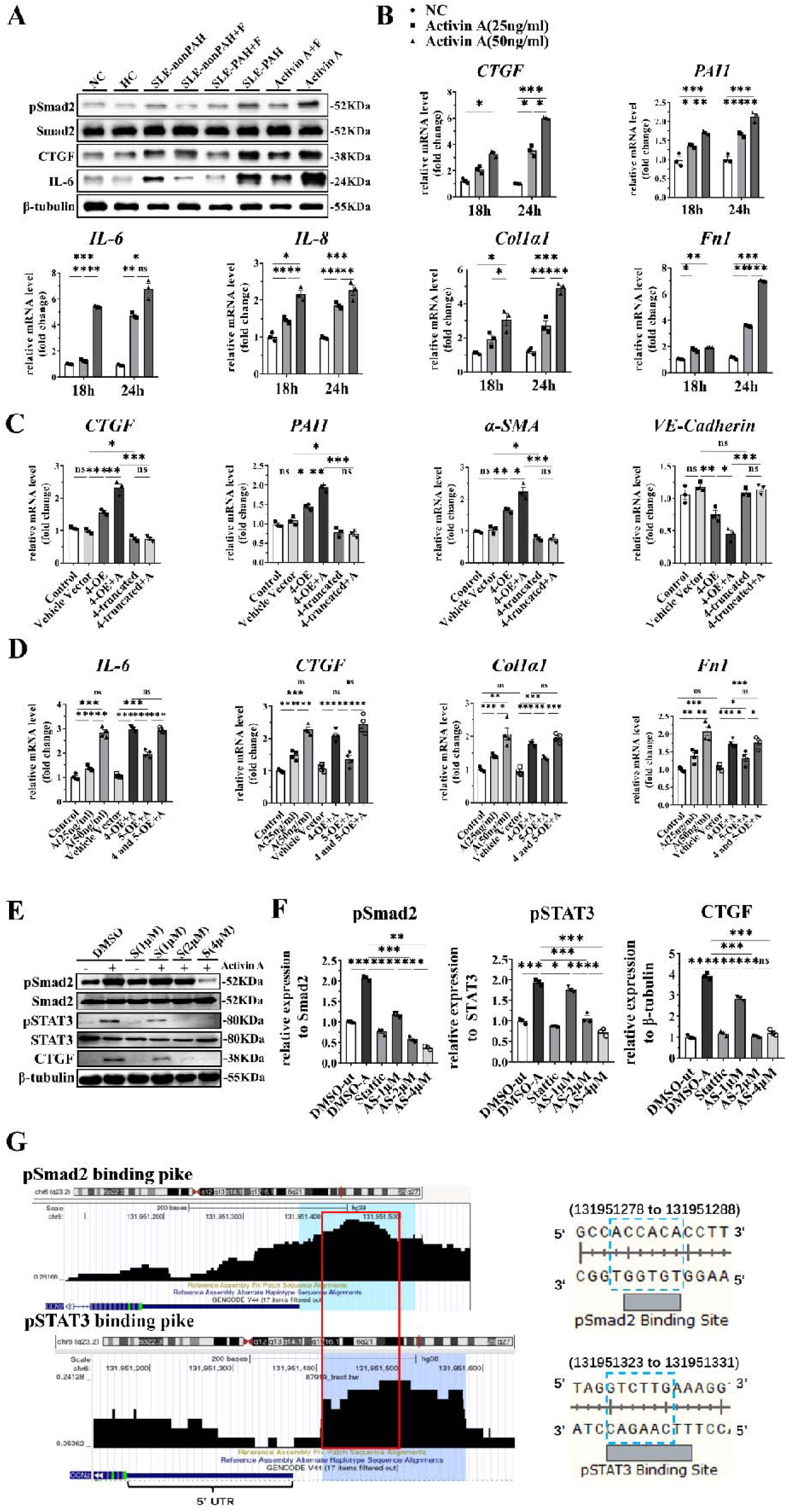
The aberrant activation of ALK4 not ALK5 induced pro-inflammatory factor and EndoMT gene expression in hPMECs. **(A)** hPMECs stimulated by HC, SLE and SLE-PAH patients’ sera, Activin A(25ng/ml) and and Follistatin (100ng/ml) for 24 h were collected and examined by western blot. NC: Negative control. **(B)** The mRNA expression in hPMECs induced by Activin A (25ng/ml and 50ng/ml) for indicated time (N=3). **(C)** The mRNA expression in Control, ALK4-OE, ALK4-truncatedand and Vehicle Vector group (N=3). **(D)** The mRNA expression in hPMECs transfected with ALK4-OE and ALK5-OE plasmid. **(E)** The pSTAT3, pSmad2 and CTGF expression in hPMECs treated with Stattic were examined by western blot. “S”: Stattic. **(F)** The densitometric quantification of protein levels of pSmad2, pSTAT3 and CTGF (N=3). **(G)** The overlapping binding region of pSmad2 and pSTAT3 in CTGF promoter region was indicated by red line (with cistrome database). OE: overexpression (N=3). “A”: Activin A, “F”: Follistatin. “ut”: untreated by Activin A, “AS”: Activin A+Stattic. Data were shown as mean ±SEM, *P* values (Bonferroni corrected) are depicted in the panels throughout all figures as \**P* < .05, \*\**P* < .01 and \*\*\**P* < .001. ns, not significant, *P* > .05. Using one-way ANOVA with repeated measures followed by Bonferroni correction for multiple comparisons. Each data point in the panels represents one independent subject.

To examine the direct effect of Activin A on hPMECs, we treated hPMECs with 25 or 50 ng/mL Activin A for 18 or 24 h (Figure 4B). We found that the expression levels of the Activin A target genes *CTGF* and *PAI1*[29], the proinflammatory factors *IL-6* and *IL-8* and ECM-related genes (e.g., *Fn1* and *Col1*α*1*) significantly increased in hPMECs in a time- and dose-dependent manner. We also detected increased expression of α-SMA and vimentin, and decreased expression of VE-Cadherin in hPMECs (Supplementary Figure 4B). The protein levels of CTGF and pSmad2 also significantly increased in hPMECs in a time-dependent manner after stimulation with Activin A (Supplementary Figure 4C and D).

Having demonstrated that ALK4 mediates IL-17 production in CD4^+^ T cells, we next assessed whether ALK4 affects hPMECs by overexpressing full-length and truncated *ALK4*. *ALK4* contains an extracellular region, transmembrane region, GS domain and protein kinase domain for phosphorylating Smad 2/3[30]. Based on the NCBI nucleotide sequence of canonical human ALK4 (NM_004302.5), we inserted a full-length or truncated *ALK4* sequence into the pcDNA3.1-Cppt-IRES plasmid for lentiviral transduction. The truncated *ALK4* sequence lacked part of the GS domain and the protein kinase domains (Supplemental Figure 5A). To elucidate the importance of ALK4 in Activin A signaling in hPMECs, we individually transfected hPMECs with the ALK4-OE, ALK4 truncation or control plasmid (Supplemental Figure 5B). Notably, ALK4 overexpression led to significant upregulation of *CTGF, PAI1*, *Col1*α*1, IL-6, IL-8* and *Fn1* indicating an aberrant increase in the expression of ECM-related genes (Figure 4C and Supplemental Figure 5C). In contrast, hPMECs transfected with the truncated ALK4 plasmid exhibited the opposite gene expression pattern, suggesting that Activin A-ALK4 signaling participates in inducing ECM-related gene expression and EndoMT in hPMECs (Figure 4C and Supplemental Figure 5C).

Both ALK4 and ALK5 are type I receptors for Activin A and TGF-β, and both are mediators of downstream signaling and function as key regulators of cell proliferation and survival[9]; however, which of these proteins plays the most crucial role in the expression of ECM-related genes, proinflammatory factors and EndoMT upon Activin A activation remains to be determined. Therefore, we stimulated hPMECs with different concentrations of Activin A and transfected hPMECs with the ALK4 and/or ALK5 plasmid. Compared with ALK5 overexpression, ALK4 overexpression induced increases in *CTGF, Fn1*, *Col1*α*1,* and *IL-6* expression, as well as mesenchymal markers, including α*-SMA* and *Vimentin* in hPMECs. However, the expression of *Bmpr2* and the endothelial marker *VE-Cadherin* was significantly decreased (Figure 4D; Supplementary Figure 5D), the protein expression of pSmad2, IL-6 and CTGF by overexpression ALK4 and ALK5 were also confirmed with immunoblotting (Supplementary Figure 5E).

Due to the key role of CTGF in promoting EndoMT and ECM-related gene expression, we further investigated the transcriptional regulation of *CTGF* in hPMECs. According to previous studies, STAT3 activation is required for signaling via Smad2-promoted CTGF transcription^32-34^. We treated hPMECs with different concentrations of Stattic (Figure 4E), which selectively inhibited Stat3 activation by blocking tyrosine phosphorylation and Stat3 dimerization^35^, and concomitantly incubated hPMECs with 25 ng/mL Activin A for 3 h. As shown in Figure 4E and Figure 4F, Stattic decreased the pSTAT3, pSmad2 and CTGF levels in hPMECs in a dose-dependent manner, indicating that pSTAT3 is an essential signaling molecule that regulates Smad2-CTGF pathway activation.

We then searched for pSmad2 and pSTAT3 binding sites in the *CTGF* promoter region using the GSM2635691 and GSM2752894 datasets and the cistrome database (cistrome.org/db/), a database compiling the genome-wide locations of transcription factor-binding sites, and JASPAR (jaspar.elixir.no), providing transcription factor binding profiles. Through further sequencing analysis, we found overlapping pSmad2- and pSTAT3-binding regions in the *CTGF* promoter, which are indicated by the blue and purple panels, respectively, on chromosome 6. The regions containing large overlaps are indicated with red rectangles in Figure 4G.

These findings highlighted the significance of ALK4 rather than ALK5 in upregulating *IL-6*, and signaling through pSTAT3 which synergistically acts with pSmad2 by binding to the CTGF promoter to drive EndoMT in hPMECs.

### 3.5. ALK4 overexpression induced severe pulmonary hypertension in SLE mice

To confirm the importance of ALK4 in vivo, we aimed to examine the detrimental role of ALK4 by establishing an SLE-PH mouse model (Figure 5A). In our previous study, mice with SLE-PH induced with pristane and exposure to hypoxic conditions (10% O_2_) (SLE-PH mice) exhibited only a mild increase in right ventricular systolic pressure (RVSP) without right heart hypotrophy. Therefore, we treated mice weekly with an ALK4-overexpressing lentivirus via intranasal instillation to reproduce the increased activation of Activin A in the SLE-PH mouse model and further monitored the mice for 5 weeks. We observed an increase in the RVSP to approximately 35 mmHg and a significant increase in the right heart hypotrophy index (Fulton index) (Figure 5B and D).

**Figure 5.**
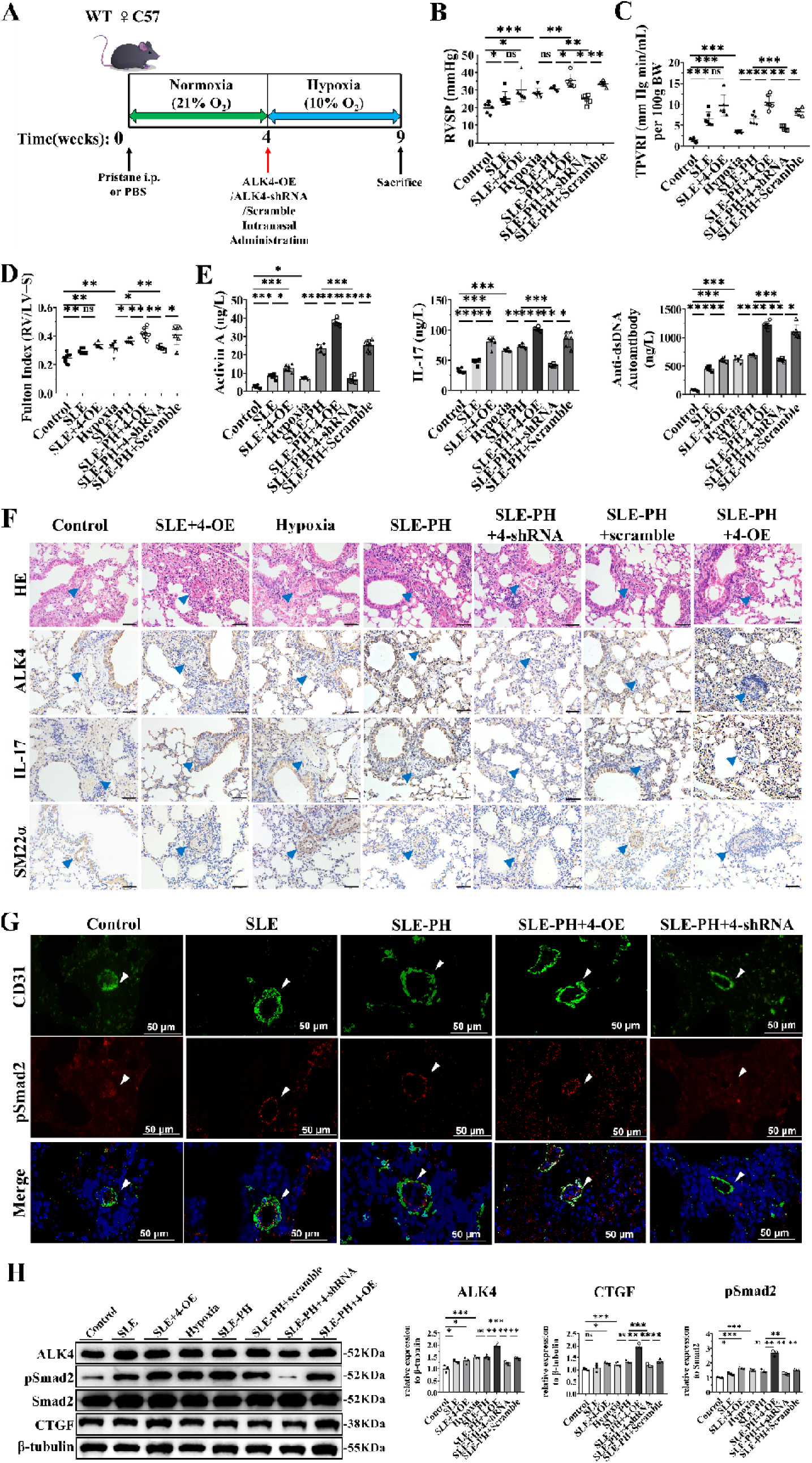
ALK4 overexpression induced severe pulmonary hypertension in SLE mice. **(A)** Schematic illustration of ALK4 overexpression and knockdown in SLE-PH mice. **(B, C, D)** Hemodynamic indices including RVSP, TPVRI and Fulton index were measured in the SLE-PH mice (N=6 each). **(E)** ELISA showing Activin A, IL-17 and anti-ds DNA autoantibody level in the serum of different groups. **(F)** The representative images of pulmonary arteries (Scale bar=50 μm) by HE and IHC staining of SM22α, ALK4 and IL-17 in each group. Blue arrows indicated small pulmonary arteries. **(G)** Representative images showed CD31 (green) and pSmad2 (red) with DAPI (blue) in SLE-PH mice. White arrows indicated the pulmonary small arteries. Scale bar=50 μm. **(H)** The western blot result showed that ALK4, pSmad2 and CTGF expression in the lungs of each group (left) with quantification (right). “4-OE”: ALK4 overexpression. “4-shRNA”: ALK4-shRNA. Data were shown as mean ±SEM, *P* values (Bonferroni corrected) are depicted in the panels throughout all figures as \**P* < .05, \*\**P* < .01 and \*\*\**P* < .001. ns, not significant. Using one-way ANOVA with repeated measures followed by Bonferroni correction for multiple comparisons.

We also applied ALK4 shRNA to confirm the reversal effects of knocking down ALK4. Compared with that in the control group, ALK4 knockdown resulted in considerable amelioration of the increase in the RVSP, a reduced total pulmonary vascular resistance index (TPVRI) and a decreased Fulton index (Figure 5B and D). The serum levels of Activin A, IL-17 and mouse anti-dsDNA autoantibodies, which are markers of SLE in humans[31] were also decreased (Figure 5E). Alleviation of pulmonary inflammation and decreased perivascular IL-17 levels were observed in the ALK4 knockdown group, accompanied by reduced infiltration of immune cells, as assessed by HE staining and IHC staining of IL-17 (Figure 5F), and the wall thickness of the small pulmonary arteries was markedly decreased, as shown by IHC staining of α-SMA (Supplemental Figure 6A). pSmad2 expression in pulmonary vasculature was examined by immunofluorescence staining, with CD31 as marker to localize PMECs (Figure 5G). The decrease in pSmad2 levels was observed in ALK4 knockdown group. A reduction in ALK4 and Smad2 phosphorylation were accompanied by reduced CTGF protein levels in the ALK4 knockdown group (Figure 5H). Further gene expression analysis revealed significant decreases in the expression levels of *IL-17*, *Rorc*, *CTGF, Col1α1, IL-6, GM-CSF* and *PAI1* in the ALK4 knockdown group, while the expression of *IL-10* was increased in this group (Supplementary Figure 6B). In addition, the expression levels of EndoMT markers (*a-SMA* and *Vimentin*) decreased and the expression of *VE-cadherin* was restored in the ALK4 knockdown group.

Our results show that ALK4 has a detrimental effect on pulmonary vascular remodeling in SLE-PH mice and that decreasing ALK4 expression reverses pulmonary hypertension in vivo.

### 3.6 The potent ALK4 inhibitor TEW reversed pulmonary hypertension by suppressing IL-17 expression in the lungs and EndoMT in hPMECs

To evaluate whether inhibition of ALK4 activation can suppress inflammation and reverse PAH in patients with SLE, we used the ALK4 inhibitor TEW, which is in a phase II clinical trial for breast cancer therapy[32, 33]. Its low IC50 (13 nM) and minimal side effects in patients make it a promising candidate for clinical application as a TGF-β receptor inhibitor[34]. Specifically, TEW exhibits a potent inhibitory effect via the inhibition of type I receptor-mediated phosphorylation of Smad2, the essential transducer of TGF-β downstream signaling[34].

Based on the efficient doses reported previously[35], we applied three different TEW doses for the in vitro assay. A significant increase in *CTGF*, *IL-6* and α*-SMA* expression was observed in hPMECs activated by IL-17 and Activin A, while the expression of the endothelial marker *VE-Cadherin* was decreased. Treatment with TEW effectively reversed the IL-17- and Activin A-induced alterations in gene expression (Figure 6A and Supplemental Figure 7A) and IL-6 rα secretion (Figure 6C) from hPMECs. IL17, GM-CSF, RORC and IL-10 expression were significantly reduced in TEW treated Th17 cells (Figure 6B and and Supplemental Figure 7B).

**Figure 6.**
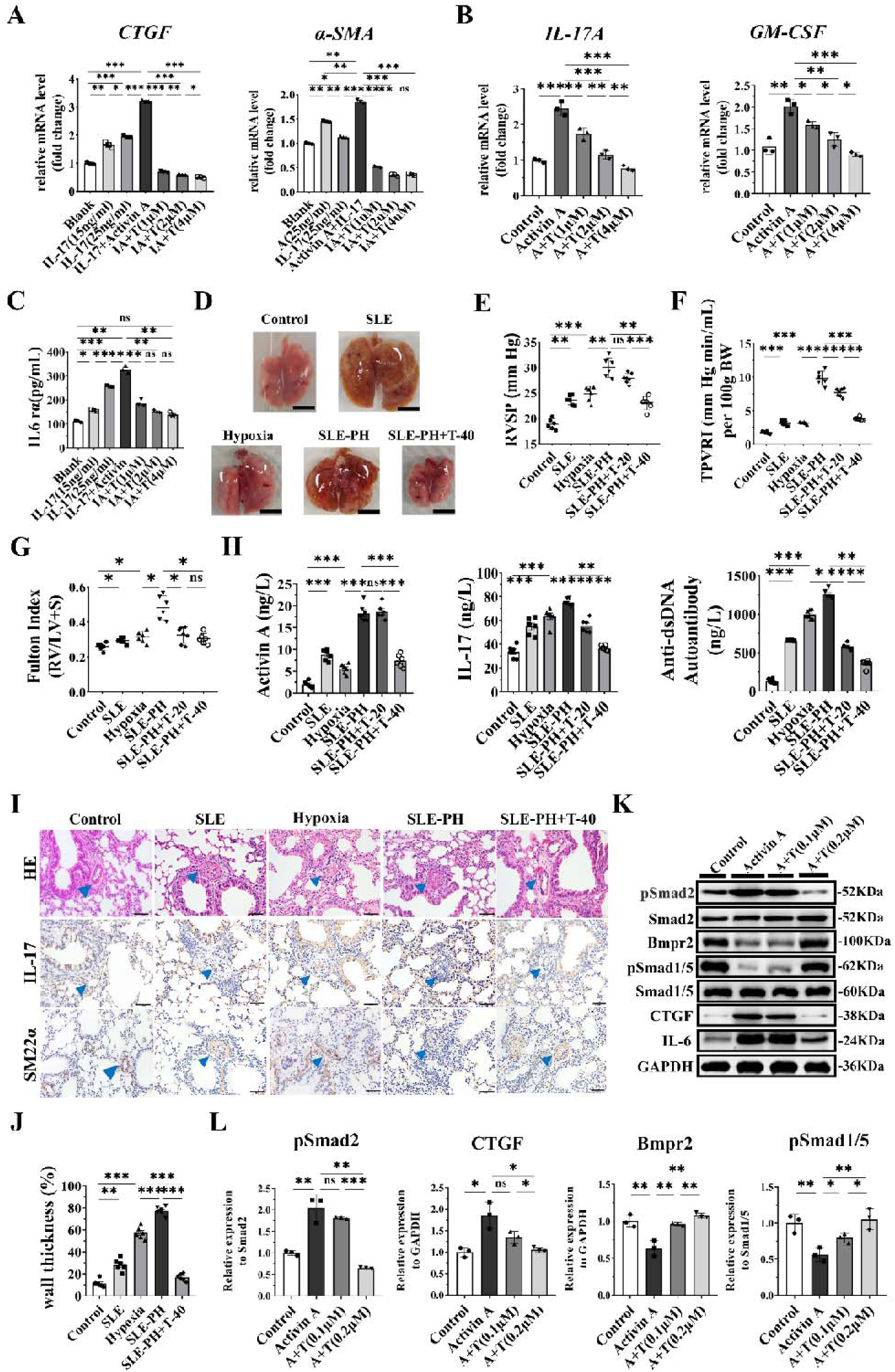
The potent ALK4 inhibitor TEW reversed pulmonary hypertension by suppressing IL-17 expression in the lungs and EndoMT in hPMECs. **(A)** The mRNA expression in hPMECs treated with TEW after stimulation with IL-17 and Activin A. “IA” represented IL-17+Activin A (N=3). **(B)** The mRNA expression in Th17 cells treated with TEW after stimulation with Activin A (N=3). **(C)** IL-6rα level in the supernatant of hPMECs treated with TEW (N=3). **(D)** Representative images of the lung of control and treated mice (Scale bars: 1cm). **(E, F, G)** Hemodynamic indices including RVSP, TPVRI and Fulton index in each group (N=6). **(H)** ELISA showing IL-17, anti-ds DNA autobody and Activin A level in the serum from different groups. **(I)** The representative images of pulmonary arteries (Scale bar=50 μm) by HE and IL-17 and SM22α IHC staining of each group. Blue arrows indicated the small pulmonary arteries. **(J)** Medial wall thickness of small pulmonary arteries. (K) The protein expressions of pSmad2, Bmpr2, pSmad1/5, CTGF and IL-6 in hPMECs treated with TEW were examined by western blot. **(K)** The densitometric quantification of protein levels of pSmad2, CTGF, Bmpr2 and pSmad1/5 (N=3). “T-20”: TEW (20mg/kg), “T-40”: TEW (40mg/kg), “AT”: Activin A +TEW. Data were shown as mean ±SEM, *P* values (Bonferroni corrected) are depicted in the panels as \**P* < .05, \*\**P* < .01 and \*\*\**P* < .001. ns, not significant. Using one-way ANOVA with repeated measures followed by Bonferroni correction for multiple comparisons.

Next, we treated SLE-PH mice with an effective dose of TEW for cancer therapy, which has been shown to rarely have side effects[32]. Images of the general lung morphology in each group after 4 weeks of treatment are shown in Figure 6C. Hemorrhage and edema were observed in the lungs of SLE and SLE-PH mice, while the mice treated with TEW had reduced edema and similar areas of hemorrhage compared to the mice in the control group (Figure 6D). Treatment with TEW effectively reversed t he increases in RVSP, TPVRI and the Fulton index in mice with SLE-PH (Figure 6E-G). Elevated serum levels of IL-17, Activin A and mouse anti-dsDNA autoantibodies were detected in the SLE and SLE-PH groups, as expected. Notably, TEW treatment significantly reduced the levels of IL-17, anti-dsDNA antibodies and Activin A (Figure 6H).

HE and IHC staining revealed significant decreases in perivascular IL-17 levels, lung inflammation and immune cell infiltration in the lungs in mice in the TEW treatment group compared with mice in the SLE-PH group (Figure 6I). The wall thickness of small pulmonary arteries also decreased markedly with TEW treatment, as evaluated by IHC staining of α-SMA (Figure 6J). Additionally, the increases in the levels of *IL-17*, *IL-6*, ECM-related genes *Col1*α*1* and *PAI1* in the SLE-PH group were significantly attenuated after treatment with TEW (Supplementary Figure 6C). We also examined α*-SMA*, *Vimentin* and *VE-cadherin* expression in the lungs of TEW-treated SLE-PH mice and showed that TEW effectively suppressed EndoMT in these mice.

We also confirmed the effects of TEW on CTGF and IL-6 protein expression and BMPR2 signaling in hPMECs. As shown in Figure 6K, 6L and Supplementary Figure 7C, TEW substantially inhibited pSmad2, CTGF and IL-6 expression in hPMECs; in contrast, the Bmpr2 and pSmad1/5 levels were significantly decreased after Activin A stimulation, but the decreases were reversed by TEW treatment.

These results demonstrated that the ALK4 inhibitor TEW effectively suppressed the expression of pro-inflammatory factors and EndoMT in hPMECs and IL-17, GM-CSF and RORC in Th17 cells. TEW contributed to the inhibition of Activin A/pSmad2 signaling while increasing BMPR2 signaling. In vivo TEW treatment reversed pathological pulmonary vascular remodeling and the development of SLE-PH.

### 3.7 TEW reversed PAH in MCT-treated rats by rebalancing signaling through the Activin A/ALK4 and BMPR2 pathways

To further confirm that TEW alleviated PAH in an inflammation-related rat model in which the animals could be catheterized more accurately, we used a rat model of MCT-induced PAH to reproduce the pathological characteristics of patients, including the infiltration of mononuclear cells in the pulmonary vasculature, that was previously reported to be used for the study of CTD-PAH[36]. Severe lung damages were observed in MCT group, while TEW treatment groups displayed less pulmonary hemorrhage (Supplementary Figure 8A and 8B). The survival rates of all groups of rats are demonstrated in Supplementary Figure 8C. TEW treatment with 40 mg/kg significantly improved the survival rate of MCT-induced rats.

Our results showed a significant improvement in RVSP, the Fulton index and the TRVRI in MCT rats treated with TEW (Supplementary Figure 8D-F). We observed that RVSP reduced to nearly 35 mmHg with lower Fulton index and TPVRI in T-40mg/kg group than MCT group. Serum IL-17 and Activin A levels were significantly elevated in MCT group and decreased in the rats administered TEW (Supplementary Figure 8G). TEW treatment also significantly reduced lung inflammation and perivascular IL-17 expression in the pulmonary vascular and significantly decrease the wall thickness of small pulmonary arteries (Supplementary Figure 9A). Additionally, the gene expression of *IL-17*, *CTGF*, *IL-6*, *Inhba*, *Rorc*, *GM-CSF*, *Col1*α*1*, *Fn1, Cxcr1* and *PAI1* decreased with TEW treatment (Supplementary Figure 8H; Supplementary Figure 9B). We also found that TEW alleviated EndoMT in MCT rats. α*-SMA* and *Vimentin* decreased and *VE-Cadherin* increased. Western blot analysis revealed a reduction in pSmad2 protein level and an elevation in Bmpr2 in the TEW-treated group (Supplementary Figure 8I). Immunofluorescence staining showed decreased pSmad2 in TEW treated Rats (Supplementary Figure 8J).

We also conducted TEW in SU5416-induced pulmonary hypertension with hypoxia in rats which blocked VEGFR-2 and caused severe pulmonary vascular remodeling (Supplementary Figure 10A). Images of the lung from each group were shown in Supplementary Figure 10B. The whole lungs showed poor blood circulation and supply in SuHx group. Our results showed a significant improvement in hemodynamic indices in the TEW treated group, which RVSP decreased from 40 to 35 mmHg with lower Fulton index and TPVRI than SuHx group (Supplementary Figure 10C-E). With TEW treatment, serum IL-17 and Activin A levels were significantly reduced (Supplementary Figure 10F); moreover, immune cell infiltration and perivascular IL-17 levels decreased (Supplementary Figure 10G).The wall thickness of pulmonary small arteries was significantly reduced with TEW treatment (Supplementary Figure 10H), as was the gene expression of *IL-17*, *IL-6*, *Inhba, Col1*α*1* and *PAI1* decreased with TEW treatment as well as mesenchymal markers (α*-SMA* and *Vimentin*) and *VE-Cadherin* increased (Supplementary Figure 10I).

The above results confirmed the cell signaling by which TEW inhibits Activin A/ALK4 signaling in immunologically dysfunctional PAH rat models.

## Discussion

This study aimed to elucidate the role of Activin A and its major downstream effector in SLE-PAH, with a special focus on the molecular targets that alleviate the initial inflammatory insult on the most vulnerable endothelial layer during the onset of PAH. We observed significantly higher levels of IL-17 and Activin A in the lungs and serum of SLE-PAH patients than in those of SLE-non PAH patients. The reduced T helper cell population in patients after treatment suggested that Th17 cells are involved in SLE-PAH. Coculture experiments revealed that ALK4 mediates the interaction between immune cells and endothelial cells. We observed increased expression of CTGF and proinflammatory genes downstream of the Activin A/ALK4 pathway in vitro and in vivo, which suggested the detrimental role of ALK4 in promoting pathogenic behaviors of Th17 cells to induce inflammation and pulmonary vascular remodeling. The specific ALK4 inhibitor TEW effectively suppressed the release of IL17 by Th17 cells and EndoMT in hPMECs in an SLE-PH mouse model and an MCT-PAH rat model. Finally, we confirmed that *CTGF* gene transcription is regulated by Activin A/pSmad2 and IL6/pSTAT3 in hPMECs. We first addressed the importance of targeting Activin A/ALK4, which also relies on suppressing pathogenic Th17 cells to strongly contribute to alleviating PAH in the context of inflammation-related disorders, such as SLE.

The potential causal mechanism is the migration of monocytes and other immune cells to the structurally damaged pulmonary artery as a result of chronic inflammation. We utilized mass cytometry to observe the dynamic involvement of immune cells before and after treatment. Mass cytometry enables the analysis of immune cells at the single-cell level, allowing the examination of the intricate immune status of patients with SLE-PAH. Following treatment, we observed reductions in the populations of T and B-cell subsets, which allowed us to identify the source of the increase in the IL-17 level.

Since the agents applied to treat SLE-PAH belong to two different groups, no therapy has been shown to achieve the ‘one stone, two birds’ effect in the clinic. The current therapeutic strategy for SLE-PAH involves combination treatment including immunosuppressive therapy along with conventional vasodilator therapy for PAH, which increases the risk of side effects[1]^1^. Immunosuppressive medications, primarily corticosteroids, cyclophosphamide and cyclosporine A, are fundamental for SLE treatment. However, these treatments fail to reverse pulmonary vascular remodeling. To improve the prognosis of SLE-PAH patients, recent efforts have been made to restore the balance between TGF-β–Activin A and BMP signaling during pulmonary hypertension progression[37]. Activin A ligand traps, such as sotatercept, were used to treat CTD-PAH patients (18%) in the STELLAR study[22, 38], suggesting the clinical effectiveness of targeting the Activin A signaling pathway in the context of CTD-PAH[21]. Our findings clarified the immunosuppressive effect in addition to the antiproliferative effect of Activin A-targeted therapy, also identifying ALK4 as the main type I receptor for Activin A in SLE-PAH.

Elevated levels of IL-6, which plays an important role in autoimmune diseases, have also been found in PAH patients[1, 39]. IL-6 is produced by macrophages and T cells through pSTAT3 to facilitate Smad2 phosphorylation upon Activin A activation. Our findings revealed that inhibiting pSTAT3 in hPMECs with Stattic led to decreased pSmad2 and CTGF levels. Thus, it is worth further exploring the interaction between the Activin A/pSmad2/CTGF pathway and IL16/pSTAT3 to determine the functional significance of the activation of pSTAT3 in hPMECs. Although we predicted the pSmad2 and pSTAT3 binding sites in the ctgf promoter region via database searches and JASPAR, the precise binding sites of pSmad2 and pSTAT3 in the ctgf promoter on chromosome 6 in hPMECs and the epigenetic modifications of this promoter remain to be confirmed. The limitations of this study include the further development of targeted therapies—for example, whether the source of Activin A in patients with SLE-PAH is the same as that in patients with PAH.

In conclusion, our research revealed that abnormally elevated Activin A levels induce IL-17 production and secretion in pathogenic Th17 cells in the context of SLE-PAH. Meanwhile, Activin A binds to ALK4 and promotes the expression of CTGF, α-SMA, and IL6 in hPMECs. Moreover, IL-6 and pSTAT3 synergistically induce CTGF expression in hPMECs. TEW effectively inhibits ALK4 activity in hPMECs and Th17 cells to alleviate inflammation and EndoMT in the context of SLE-PAH and PAH associated with other inflammatory conditions (Summary Diagram). Furthermore, we intend to utilize Activin A as a biomarker to identify the most suitable patient cohort and explore the potential development of TEW-like compounds for therapeutic application.

## Summary Diagram

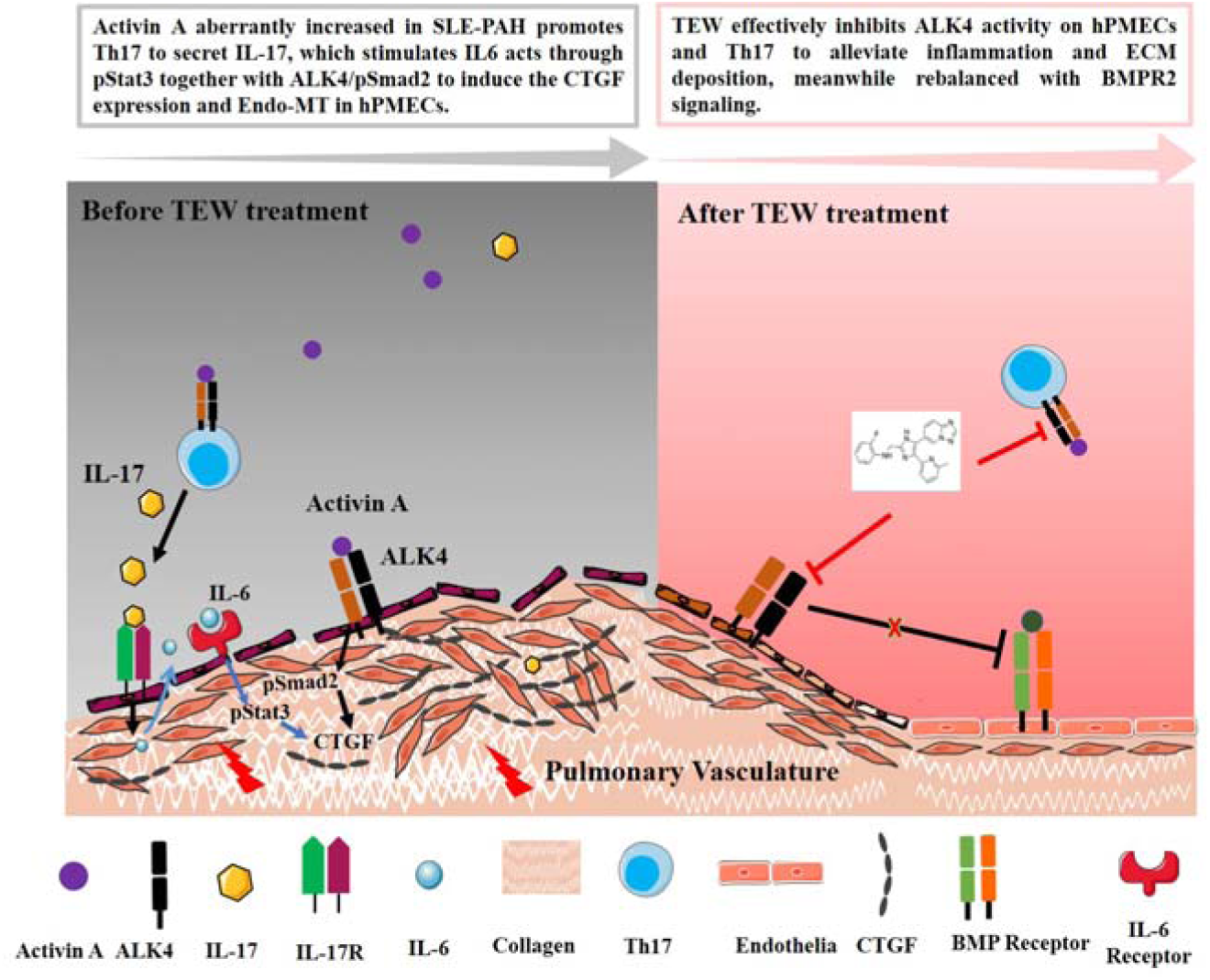

## Acknowledgement

We would like to gratefully thank the participation of patients, Hangzhou Leading Pharmatech Co., Ltd., and the Centre for Experimental Animal Research and Facility for the core facility platform of Zhejiang University School of Medicine for providing technical support.

## Contributors

Jun Yang designed the study and finalized the manuscript. Shuliang Jing and Junyan Qian, Hongjie Ying conducted the experiments and acquired data. Pei Mao and Muhammad Saleeem Iqbal Khan provided cellular molecular technique support. Yufang Ding, has collected patient samples. Mengtao Li provided the patient information.

## Patient consent for publication

Not applicable.

## Ethics approval

All individuals provided written informed permission by the principles established in the declaration of Helsinki approved by the Ethics Committee of Zhejiang University Medical College (No.014-2015). All animal study protocols were reviewed and approved by Animal Care & Use Committee of Zhejiang University (ZJU20210036 and ZJU20240927, AIRB-2023-0317).

## Data availability statement

The data that support the findings of this study are available on request from the corresponding author upon reasonable request. All data relevant to the study are included in the article or uploaded as supplementary information.

## Reference

1. Zhao J, Wang Q, Deng X, Qian J, Tian Z, Liu Y, Li M, Zeng X: The treatment strategy of connective tissue disease associated pulmonary arterial hypertension: Evolving into the future. Pharmacology & Therapeutics 2022, 239.

2. Chaisson NF, Hassoun PM: Systemic Sclerosis-Associated Pulmonary Arterial Hypertension. Chest 2013, 144(4):1346–1356.

3. Nihtyanova SI, Schreiber BE, Ong VH, Rosenberg D, Moinzadeh P, Coghlan JG, Wells AU, Denton CP: Prediction of Pulmonary Complications and Long-Term Survival in Systemic Sclerosis. Arthritis & Rheumatology 2014, 66(6):1625–1635.

4. Gaowa S, Zhou W, Jiang H: Effect of Th17 and Treg Axis Disorder on Outcomes of Pulmonary Arterial Hypertension in Connective Tissue Diseases. Journal of the American College of Cardiology 2014, 64(16):C218–C218.

5. Maston LD, Jones DT, Giermakowska W, Howard TA, Cannon JL, Wang W, Wei Y, Xuan W, Resta TC, Gonzalez Bosc LV: Central role of T helper 17 cells in chronic hypoxia-induced pulmonary hypertension. American journal of physiology Lung cellular and molecular physiology 2017, 312(5):L609–L624.

6. Singh RP, Hasan S, Sharma S, Nagra S, Yamaguchi DT, Wong DTW, Hahn BH, Hossain A: Th17 cells in inflammation and autoimmunity. Autoimmunity Reviews 2014, 13(12):1174–1181.

7. Wu B, Zhang S, Guo Z, Bi Y, Zhou M, Li P, Seyedsadr M, Xu X, Li J-l, Markovic-Plese S et al: The TGF-β superfamily cytokine Activin-A is induced during autoimmune neuroinflammation and drives pathogenic Th17 cell differentiation. Immunity 2021, 54(2):308–323.

8. Park H, Li ZX, Yang XO, Chang SH, Nurieva R, Wang YH, Wang Y, Hood L, Zhu Z, Tian Q et al: A distinct lineage of CD4 T cells regulates tissue inflammation by producing interleukin 17. Nature Immunology 2005, 6(11):1133–1141.

9. Derynck R, Budi EH: Specificity, versatility, and control of TGF-β family signaling. Science Signaling 2019, 12(570).

10. Parichatikanond W, Luangmonkong T, Mangmool S, Kurose H: Therapeutic Targets for the Treatment of Cardiac Fibrosis and Cancer: Focusing on TGF-β Signaling. Frontiers in Cardiovascular Medicine 2020, 7.

11. Cuthbertson I, Morrell NW, Caruso P: BMPR2 Mutation and Metabolic Reprogramming in Pulmonary Arterial Hypertension. Circulation Research 2023, 132(1):109–126.

12. Aldred MA, Morrell NW, Guignabert C: New Mutations and Pathogenesis of Pulmonary Hypertension: Progress and Puzzles in Disease Pathogenesis. Circulation Research 2022, 130(9):1365–1381.

13. Ryanto GRT, Ikeda K, Miyagawa K, Tu L, Guignabert C, Humbert M, Fujiyama T, Yanagisawa M, Hirata K-i, Emoto N: An endothelial activin A-bone morphogenetic protein receptor type 2 link is overdriven in pulmonary hypertension. Nature Communications 2021, 12(1).

14. Bloise E, Ciarmela P, Dela Cruz C, Luisi S, Petraglia F, Reis FM: ACTIVIN A IN MAMMALIAN PHYSIOLOGY. Physiological Reviews 2019, 99(1):739–780.

15. de Kretser DM, O’Hehir RE, Hardy CL, Hedger MP: The roles of activin A and its binding protein, follistatin, in inflammation and tissue repair. Molecular and Cellular Endocrinology 2012, 359(1-2):101–106.

16. Tam AYY, Horwell AL, Trinder SL, Khan K, Xu S, Ong V, Denton CP, Norman JT, Holmes AM, Bou-Gharios G et al: Selective deletion of connective tissue growth factor attenuates experimentally-induced pulmonary fibrosis and pulmonary arterial hypertension. International Journal of Biochemistry & Cell Biology 2021, 134.

17. Xing Y, Zhao S, Wei Q, Gong S, Zhao X, Zhou F, Ai-Lamki R, Ortmann D, Du M, Pedersen R et al: A novel piperidine identified by stem cell-based screening attenuates pulmonary arterial hypertension by regulating BMP2 and PTGS2 levels. European Respiratory Journal 2018, 51(4).

18. Guignabert C, Savale L, Boucly A, Thuillet R, Tu L, Ottaviani M, Rhodes CJ, De Groote P, Prevot G, Bergot E et al: Serum and Pulmonary Expression Profiles of the Activin Signaling System in Pulmonary Arterial Hypertension. Circulation 2023, 147(24):1809–1822.

19. Nihtyanova SI, Denton CP: Pathogenesis of systemic sclerosis associated interstitial lung disease. Journal of Scleroderma and Related Disorders 2020, 5(2_SUPPL):6-16.

20. Yndestad A, Larsen K-O, Oie E, Ueland T, Smith C, Halvorsen B, Sjaastad I, Skjonsberg OH, Pedersen TM, Anfinsen O-G et al: Elevated levels of activin A in clinical and experimental pulmonary hypertension. Journal of Applied Physiology 2009, 106(4):1356–1364.

21. Humbert M, McLaughlin V, Gibbs JSR, Gomberg-Maitland M, Hoeper MM, Preston IR, Souza R, Waxman A, Escribano Subias P, Feldman J et al: Sotatercept for the Treatment of Pulmonary Arterial Hypertension. New England Journal of Medicine 2021, 384(13):1204–1215.

22. Hoeper MM, Badesch DB, Ghofrani HA, Gibbs JSR, Gomberg-Maitland M, McLaughlin VV, Preston IR, Souza R, Waxman AB, Gruenig E et al: Phase 3 Trial of Sotatercept for Treatment of Pulmonary Arterial Hypertension. New England Journal of Medicine 2023, 388(16):1478–1490.

23. Galiè N, Humbert M, Vachiery J-L, Gibbs S, Lang I, Torbicki A, Simonneau G, Peacock A, Vonk Noordegraaf A, Beghetti M et al: 2015 ESC/ERS Guidelines for the diagnosis and treatment of pulmonary hypertension. European Heart Journal 2016, 37(1):67-119.

24. Hochberg MC: Updating the American College of Rheumatology revised criteria for the classification of systemic lupus erythematosus. Arthritis and Rheumatism 1997, 40(9):1725-1725.

25. du Sert NP, Hurst V, Ahluwalia A, Alam S, Avey MT, Baker M, Browne WJ, Clark A, Cuthill IC, Dirnagl U et al: The ARRIVE guidelines 2.0: Updated guidelines for reporting animal research. Plos Biology 2020, 18(7).

26. Bedoya SK, Lam B, Lau K, Larkin J, III: Th17 Cells in Immunity and Autoimmunity. Clinical & Developmental Immunology 2013.

27. Hedger MP, de Kretser DM: The activins and their binding protein, follistatin-Diagnostic and therapeutic targets in inflammatory disease and fibrosis. Cytokine Growth Factor Rev 2013, 24(3):285–295.

28. He M, Chen Z, Martin M, Zhang J, Sangwung P, Woo B, Tremoulet AH, Shimizu C, Jain MK, Burns JC et al: miR-483 Targeting of CTGF Suppresses Endothelial-to-Mesenchymal Transition Therapeutic Implications in Kawasaki Disease. Circulation Research 2017, 120(2):354-+.

29. Chen B, Chang H-M, Zhang Z, Cao Y, Leung PCK: ALK4-SMAD3/4 mediates the effects of activin A on the upregulation of PAI-1 in human granulosa lutein cells. Molecular and Cellular Endocrinology 2020, 505.

30. Attisano L, Wrana JL, Montalvo E, Massague J: Activation of signalling by the activin receptor complex. Molecular and Cellular Biology 1996, 16(3):1066–1073.

31. Freitas EC, de Oliveira MS, Monticielo OA: Pristane-induced lupus: considerations on this experimental model. Clinical Rheumatology 2017, 36(11):2403–2414.

32. Son JY, Park S-Y, Kim S-J, Lee SJ, Park S-A, Kim M-J, Kim SW, Kim D-K, Nam J-S, Sheen YY: EW-7197, a Novel ALK-5 Kinase Inhibitor, Potently Inhibits Breast to Lung Metastasis. Molecular Cancer Therapeutics 2014, 13(7):1704-1716.

33. Jung SY, Yug JS, Clarke JM, Bauer TM, Keedy VL, Hwang S, Kim S-J, Chung EK, Lee JI: Population pharmacokinetics of vactosertib, a new TGF-β receptor type Ι inhibitor, in patients with advanced solid tumors. Cancer Chemotherapy and Pharmacology 2020, 85(1):173–183.

34. Jin CH, Krishnaiah M, Sreenu D, Subrahmanyam VB, Rao KS, Lee HJ, Park S-J, Park H-J, Lee K, Sheen YY et al: Discovery of N-((4-(1,2,4 Triazolo 1,5-a pyridin-6-yl)-5-(6-methylpyri din-2-yl)-1H-imidazol-2-yl)methyl)-2-fluoroaniline (EW-7197): A Highly Potent, Selective, and Orally Bioavailable Inhibitor of TGF-β Type I Receptor Kinase as Cancer Immunotherapeutic/Antifibrotic Agent. Journal of Medicinal Chemistry 2014, 57(10):4213–4238.

35. Hong E, Park S, Ooshima A, Hong CP, Park J, Heo JS, Lee S, An H, Kang JM, Park SH et al: Inhibition of TGF-β signalling in combination with nal-IRI plus 5-Fluorouracil/Leucovorin suppresses invasion and prolongs survival in pancreatic tumour mouse models. Scientific Reports 2020, 10(1).

36. Nogueira-Ferreira R, Vitorino R, Ferreira R, Henriques-Coelho T: Exploring the monocrotaline animal model for the study of pulmonary arterial hypertension: A network approach. Pulmonary Pharmacology & Therapeutics 2015, 35:8–16.

37. Westerhof BE, Saouti N, van der Laarse WJ, Westerhof N, Noordegraaf AV: Treatment strategies for the right heart in pulmonary hypertension. Cardiovascular Research 2017, 113(12):1465–1473.

38. Souza R, Badesch DB, Ghofrani HA, Gibbs JSR, Gomberg-Maitland M, McLaughlin VV, Preston IR, Waxman AB, Gruenig E, Kope G et al: Effects of sotatercept on haemodynamics and right heart function: analysis of the STELLAR trial. European Respiratory Journal 2023, 62(3).

39. Florentin J, Zhao J, Tai Y-Y, Vasamsetti SB, O’Neil SP, Kumar R, Arunkumar A, Watson A, Sembrat J, Bullock GC et al: Interleukin-6 mediates neutrophil mobilization from bone marrow in pulmonary hypertension. Cellular & Molecular Immunology 2021, 18(2):374–384.

