## Supplemental methods and figure 1 to figure 10 for "Activin A-activated ALK4 induces pathogenic Th17 involvement in endothelial–mesenchymal transition in systemic lupus erythematosus-associated pulmonary arterial hypertension"

**Supplemental Material**

### Materials and Methods

**Mass cytometry**

Peripheral blood (2 ml) was collected from 4 patients (female) with SLE-PAH before first treatment of glucocorticoids and vasodilators and after 6 months of therapy. The peripheral blood monocytes (PBMCs) were treated according to the MaxPar Cell Surface Staining Protocol provided by the manufacturer and stained with the MaxPar Human Peripheral Blood Phenotyping Panel Kit (Fluidigm, South San Francisco, CA). Data collection was performed on a CyTOF mass cytometer. The PBMCs were isolated by using the instruction of kit (TBD Science, Tianjin, China).

**ALK4 recombinant plasmid construction**

ALK4 was identified as a transmembrane protein comprising GS (amino acids 177 to 206) and protein kinase domains with phosphorylation sites (amino acids 207 to 497) for Smad 2/3(1). Based on the NCBI nucleotide sequence of canonical human ALK4 (NM_004302.5), we constructed a recombinant plasmid containing the full-length nucleotide sequence of ALK4 (1517 bp, from 5’ to 3’) and a recombinant plasmid containing a truncated ALK4 sequence (553 bp, from 5’ to 3’) that included the extracellular region, transmembrane domain and part of the GS domain but did not include the protein kinase domains. The ALK4 and ALK5 full-length overexpression plasmids were purchased from Vigene Bioscience (Shandong, China). We constructed a vector for stable expression of full-length ALK4, a vector for stable expression of a truncated ALK4 protein and a vehicle vector for lentiviral transduction into splenocytes and hPMECs. For the production of the lentiviral supernatant, the day before transduction, HEK293T cells were seeded at 8 × 10^6^ cells per 100 mm dish. The lentiviruses were produced by co-transfecting HEK293T cells with a plasmid containing the ALK4 gene sequence (13.5 μg), the pMD.2 G plasmid encoding the VSV-G envelope protein (7.5 μg), and the packaging plasmid psPAX2 (16.5 μg) using PEI for 24 h following the manufacturer’s instructions. The supernatants were collected after 48 h and filtered through a 0.45 μm membrane to remove cell debris. The lentiviruses were concentrated by ultracentrifugation (Optima XE-100, Beckmann) at 827,000 × g for 2 h at 4 °C.

**In vitro assays**

hPMECs lines were bought from Lonza and cultured in an ECM medium (5% FBS, 1% Penicillin-Streptomycin Solution and 1% Endothelial Cell Growth Supplement, Sciencell, USA). Human recombinant Activin A (25 ng/mL or 50 ng/mL, SinoBiological, China) and Human recombinant IL-17 (25 ng/mL, SinoBiological, China) were added to the hPMEC medium for 12 h at 37°C with 1 μM, 2 μM and 4 μM TEW after serum-starved with 1% FBS for 12 hours. After 24 h, the cells were collected for further assays. For mechanism assay, after hPMECs being serum-starved with 1% FBS for 12 hours, Stat3 inhibitor static (Selleck, USA) was added into fresh ECM medium for hPMECs cultured in 12-well plate and then activated with 25 ng/ml Activin A. After 4 h, the cells were collected for further assays. For TEW inhibition mechanism assay, after serum-starved with 1% FBS for 12 hours, 0.1 μM and 0.2 μM TEW (Selleck, USA) were added into fresh ECM medium for hPMECs cultured in 12-well plate activated by 25 ng/ml Activin A. After 12 h, the cells were collected for further assays.

**Quantitative real-time PCR**

Total cellular RNA was extracted using the TRIzol reagent (Invitrogen, Carlsbad, USA). According to the manufacturer’s instructions, complementary DNA (cDNA) templates were generated from 2 μg of total RNA using the PrimeScript™ RT reagent kit with gDNA Eraser (Takara Bio, Kusatsu, Japan). Quantitative real-time PCR (qPCR) was performed using the TransStart Tip Green qPCR SuperMix (TransGen Biotech, Beijing, China). The CFX Connect™ Real-Time PCR Detection System (Bio-Rad, Foster City, USA) and CFX Manager software were used to analyze the expression levels of target genes. relative gene expression = 2−ΔΔCT. The employed qPCR primers used for qPCR analysis are listed below.

| Gene | Species | Sequences |
| --- | --- | --- |
| *IL-6* | human | Forward Primer: atgaactccttctccacaagcgcct  Reverse Primer: caaatctgttctggaggtactctag |
| *Bmpr2* | human | Forward Primer: atgacttcctcgctgcagcggccct  Reverse Primer: atccttgttttacaagatttatgtc |
| *CTGF* | human | Forward Primer: aagggcaaaaagtgcatccgtactc  Reverse Primer: tcatgccatgtctccgtacatcttc |
| *VE-Cadherin* | human | Forward Primer: atgcagaggctcatgatgctcctcg  Reverse Primer: agtctccaggttttcgccagtgtcc |
| *α-SMA* | human | Forward Primer: atgtgtgaagaagaggacagcactg  Reverse Primer: tggctgggacattgaaagtctcaaa |
| *Col1α1* | human | Forward Primer: atgttcagctttgtggacctccggc  Reverse Primer: acgccggtggtttcttggtcggtgg |
| *Fn1* | human | Forward Primer: agcggacctacctaggcaatgcgtt  Reverse Primer: catggcagcggtttgcgatggtaca |
| *Vimentin* | human | Forward Primer: acgtgactacgtccacccgcaccta  Reverse Primer: agctccaccttctcgttggtgcggg |
| *PAI1* | human | Forward Primer: atgcagatgtctccagccctcacct  Reverse Primer: catgaagccctggaccagcttcaga |
| *ALK4* | human | Forward Primer: acgctccaggatcttgtctacgatc |
|  |  | Reverse Primer: gacaaggccagcttaatcatcccct |
| *IL-8* | human | Forward Primer: atgacttccaagctggccgtggctc |
|  |  | Reverse Primer: gttttccttggggtccagacagagc |
| *IL-17* | Rat | Forward Primer: atgagtccccggagaattccatcca  Reverse Primer: aggggcacttctcaggctccctctt |
| *CTGF* | Rat | Forward Primer: atgctcgcctccgtcgcgggtcccg  Reverse Primer: ggggcacacagcccacggccccatc |
| *Rorc* | Rat | Forward Primer: atggaagtcgtcctcgtcagaatgt  Reverse Primer: gtggaggtgctggaagtcctgtagc |
| *Col1α1* | Rat | Forward Primer; atgttcagctttgtggacctccggc  Reverse Primer: aggaccaggaagtccaggctgtcca |
| *IL-6* | Rat | Forward Primer: atgaagtttctctccgcaagagact  Reverse Primer: tatcttgtaagttgttcttcacaaa |
| *GM-CSF* | Rat | Forward Primer: acccaaccctgtcacccggccctg  Reverse Primer: acttctatttcacagtcagtttccg |
| *INHBA (Activin A)* | Rat | Forward Primer: atgcccttgctttggctgagaggat  Reverse Primer: cctggctgtgcctgactcggcaaag |
| *α-SMA* | Rat | Forward Primer: ctctggtgtgtgacaatggctccgg  Reverse Primer: cataatctgggtcattttctcccgg |
| *Vimentin* | Rat | Forward Primer: tgtccaccaggtccgtgtcctcgtc  Reverse Primer: ctcctgcaattccaccttctcgttg |
| *VE-Cadherin* | Rat | Forward Primer: atgcagaggctcacagagctggcca  Reverse Primer: agtaaggaagtactcagagactttt |
| *PAI1* | Rat | Forward Primer: atgcagatgtcttcagccctcactt  Reverse Primer: atgaccccatgagctccttggagag |
| *IL-6* | Rat | Forward Primer: atgaagtttctctccgcaagagact  Reverse Primer: tatcttgtaagttgttcttcacaaa |
| *Cxcr1* | Rat | Forward Primer: atggccgaggctgagtatttcatct  Reverse Primer: cccttccatttggagacagccaaga |
| *Fn1* | Rat | Forward Primer: tgctgctgctagcagtcctgtgcct  Reverse Primer: gtgtcacccactttgtaagtgtttc |
| *IL-6* | Mouse | Forward Primer: atgaagttcctctctgcaagagact  Reverse Primer: ctaggtttgccgagtagatctcaaa |
| *IL-17* | Mouse | Forward Primer: atgagtccagggagagcttcatct  Reverse Primer: ttaggctgcctggcggacaatcga |
| *α-SMA* | Mouse | Forward Primer: gccctggtgtgcgacaatggctctg |
|  |  | Reverse Primer: agtggtgcctctgtcagcagtgtcg |
| *Vimentin* | Mouse | Forward Primer: atgtctaccaggtctgtgtcctcgt  Reverse Primer: ctgcagttctaccttctcgttggtg |
| *VE-Cadherin* | Mouse | Forward Primer: atgcagaggctcacagagctggcca  Reverse Primer: tcccggtctagcctctcataggcaa |
| *PAI1* | Mouse | Forward Primer: atgcagatgtcttcagcccttgctt |
|  |  | Reverse Primer: cacggccccatgagctccttggaga |
| *INHBA (Activin A)* | Mouse | Forward Primer: atgcccttgctttggctgagaggat  Reverse Primer: agctttctgatcgcgttgagaagcg |
| *CTGF* | Mouse | Forward Primer: atgctcgcctccgtcgcaggtccca  Reverse Primer: gcacgcagcccacggccccatcca |
| *Rorc* | Mouse | Forward Primer: atggacagggccccacagagacacc  Reverse Primer: tcttggccacttgttcctgttgctg |
| *Fn1* | Mouse | Forward Primer: atgctcaggggtccgggacccggg  Reverse Primer: cctgcctctcccagccccgatgcag |

**Western blotting**

Different groups of cells were grown to confluence in 6-well plates and maintained in ECM medium. 50 μg of lysates of all were loaded onto SDS-PAGE gels and transferred to PVDF membranes using a semi-dry transfer system (Bio-Rad). The primary antibodies used in the study were anti-GAPDH (TransGen, HC301-01), anti-IL-6 (Beyotime, AF0201), anti-βtubulin(TransGen, HC101-01), anti-CTGF (Abclonal, A11067), anti-smad1/5 (Abclonal, A23208), anti-smad2 (Abclonal, A11498), anti-ERK1/2 (abclonal, A4782), anti-pERK1/2 (Beyotime, AF1891), anti-pSmad2 (Abclonal, AP0548), anti-pSmad1/5 (Cell Signaling Technology, 9516T), anti-BMPR2 (Abclonal, A16778), anti-Stat3 (Beyotime, AF1492) and anti-pStat3 (Beyotime, AF5941). The samples derive from the same experiment and that blots were processed in parallel.

**Immunofluorescence Staining**

For mouse pulmonary endothelial cells, anti-SM22α antibody (Santa, sc-373928) was used for staining following formalin-fixed and washing with PBS. The secondary antibodies were labelled with anti-mouse Alexa Fluor594 (Invitrogen, R37115). Nuclei were stained with DAPI reagent, prior to imaging on an Olympus confocal microscope (FV1000MPE, Tokyo, Japan). Five randomly selected fields from each group were observed in an unbiased fashion.

**WT C57BL/6J** **mouse pulmonary endothelial cells isolation**

Briefly, WT C57BL/6J mouse lung was isolated and the margin part tissue of lung was cut for extracting pulmonary endothelial cells. After washing off blood with hank’s solution, the tissue was further cut into small size (about 1-2mm) in tubes and seeded into 6 cm culture dishes with fresh ECM medium (5% FBS, 1% Penicillin-Streptomycin Solution and 1% Endothelial Cell Growth Supplement, Sciencell, USA). The lung tissue was removed after 60 hours and the pulmonary endothelial cells were grown to confluency.

**Th17 cells and mouse pulmonary endothelial cells co-culture assay**

In vitro non-contact co-culture assay was used to investigate the interaction between Th17 cells and mouse pulmonary endothelial cells. A mount of 5 × 10^5^ mouse pulmonary endothelial cells per well was incubated into 24-well cell culture plate. A mount of 1 × 10^5^ cells of Th17 cells per well was seeded into transwell clear inserts (Corning, USA). After co-culture for 36 hours, Th17 cells in transwell clear inserts were removed and mouse pulmonary endothelial cells were collected for immunofluorescence staining and quantitative real-time PCR.

**WT C57BL/6J** **mouse splenocyte isolation and ELISA assay**

WT C57BL/6J mouse spleen was isolated and ground into cell suspension after filtering by 25 μm filter and red cell lysis was used to erase red blood cells. After centrifugating at 200g, 4℃, 10 minutes, the splenocytes were washed by PBS two times and cultured in 1640 medium with 10% FBS and 1% Penicillin-Streptomycin Solution at 37°C in the incubator activated with 5 μg/mL PHA (Solarbio, P8090).

The levels of IL-17 and Activin A, serum of the control mouse, SLE, hypoxia, SLE-PH and SLE-PH-TEW mice, as well as serum from healthy controls and patients were assayed using IL-17 (Hengrui Hongchuang Technology Development Co., China) and Activin A (Hengrui Hongchuang Technology Development Co., China) ELISA kits according to the manufacturer's instructions. The levels of dsDNA auto-Abs (Bluewood) in control, SLE, hypoxia, SLE-PAH and SLE-PAH-TEW mice were assayed using dsDNA auto-Abs (Hengrui Hongchuang Technology Development Co., China) ELISA kits according to the manufacturer’s instructions. After hPAECs were treated with TEW after stimulation with IL-17 and Activin A for 48 h, the cell media was collected for IL-6rα ELISA, according to the manufacturer’s instructions. The absorbance was read at 450 nm, and the results are presented as endotoxin units (EU) per mL.

**In vitro T cell culture, activation and differentiation**

1x10^6^ Naïve CD4^+^ T cells were isolated from the WT mouse spleen using Mo joSort Mouse CD4 Naive T Cell Isolation Kit (480039-BLG, Miltenyi Biotec) and activated in 6 well-plates pre-coated with 3 µg/mL anti-CD3 (14-0032-81, ThermoFisher) and 5 µg/mL anti-CD28 mAb (14-0281-81, ThermoFisher) in RPMI1640 medium (Lonza). For Activin-A+IL-6 promoted Th17 cells, 30 ng/ml Activin-A (Sinobiological, 10429-HNAH) and 40 ng/ml IL-6 (MCE, HY-P7063) were added to the culture for 4 days.

**CD4^+^ T cells depletion**

CD4^+^ T cell depletion was performed by intraperitoneal administration of monoclonal anti-CD4 antibody (clone GK1.5, Bio X Cell) for a single dose per mouse once a week. Each mouse received 400 μg of GK1.5 or control rat isotype antibody IgG2b (Abclonal) in a volume of 200 μl PBS.

**ALK4 overexpression, ALK4 knockdown and TEW treatments in SLE-PH mouse**

For ALK4 expression and ALK4 knockdown treatments, after four weeks in normoxia, ALK4 overexpression lentivirus (1×10E10 PFU), ALK4-shRNA (1×10E10 PFU) lentivirus and scramble-shRNA (1×10E10 PFU) lentivirus were nasally dropped in corresponding groups every week lasting 4 weeks.

For TEW treatment, 20mg/kg and 40mg/kg TEW (TEW, Selleck, USA) were orally administered to the SLE-PH+TEW group every two days starting at the 6th week until the endpoint of model establishment. SLE-PH group was received comparable saline.

**MCT and Sugen-hypoxia Rat Pulmonary Hypertension Models**

Adult male Sprague-Dawley rats (170-190 g) were used for all animal experiments, which the Animal Research Committee approved. For the MCT model, rats were randomly injected intraperitoneally with MCT (55 mg/kg), followed by 21 days of normoxia. TEW (20mg/kg and 40mg/kg) was orally administered to rats on days 21, 28 and 32 and sacrificed on the 35th day. Vehicle was received comparable saline. Age-matched control rats were maintained under normoxia conditions for 28 days. For the Sugen-hypoxia PH model, rats were subcutaneously randomly injected with Sugen 5416 (30 mg/kg, Aladdin, China) followed by hypoxia at 10% O2 for 21 days, and 14 days of normoxia. TEW (40 mg/kg) was orally administered to the rats every other day starting from the 14th day. Vevo 2100 system (Visualsonics, Toronto, ON, Canada) was used to perform transthoracic echocardiography according to manufacturer’s instructions.

To perform hemodynamic measurements, rats were anaesthetized with pentobarbital sodium (2% in 0.9% saline, 0.2 mL/100 g body weight) injected intraperitoneally once, mice were anaesthetized with pentobarbital sodium (1% in 0.9% saline, 0.1 mL/10 g body weight) injected intraperitoneally once. The Fulton index was calculated as the weight of the right ventricle divided by the weight of the left ventricle and the weight of the septum. The BL-420S physiological experimental system was used to monitor RVSP. The right lung was embedded in a 10% neutral buffered formalin solution for histological analysis. Serum was collected by centrifugation (1000 rpm for 15 min).

**Hematoxylin–Eosin and Immunohistochemical staining**

The right lungs and heart tissues were immobilized in a 4% paraformaldehyde solution and embedded in paraffin. Hematoxylin–eosin staining was performed as described in the previous article(2). Morphological changes and degree of damage were detected with hematoxylin–eosin staining for light microscopy. The wall thickness of pulmonary arteries was calculated with diameter 50–200 µm, % = medial wall thickness/outer radius.

Part of the right lungs were immobilized in a 4% paraformaldehyde solution and embedded in paraffin. After the lungs were deparaffinized, they were stained for immunohistochemical staining with anti-ALK4 (Abclonal, A2279) and anti-IL17 (Abclonal, A21266). Immunohistochemical (IHC) staining was performed as previously described. The secondary antibody was anti-rabbit AlexaFluor488 (Invitrogen, A-11008). Images were obtained using a Carl Zeiss Axio Vision microscope (Carl Zeiss, Jena, Germany).

### Supplemental Figures and Tables

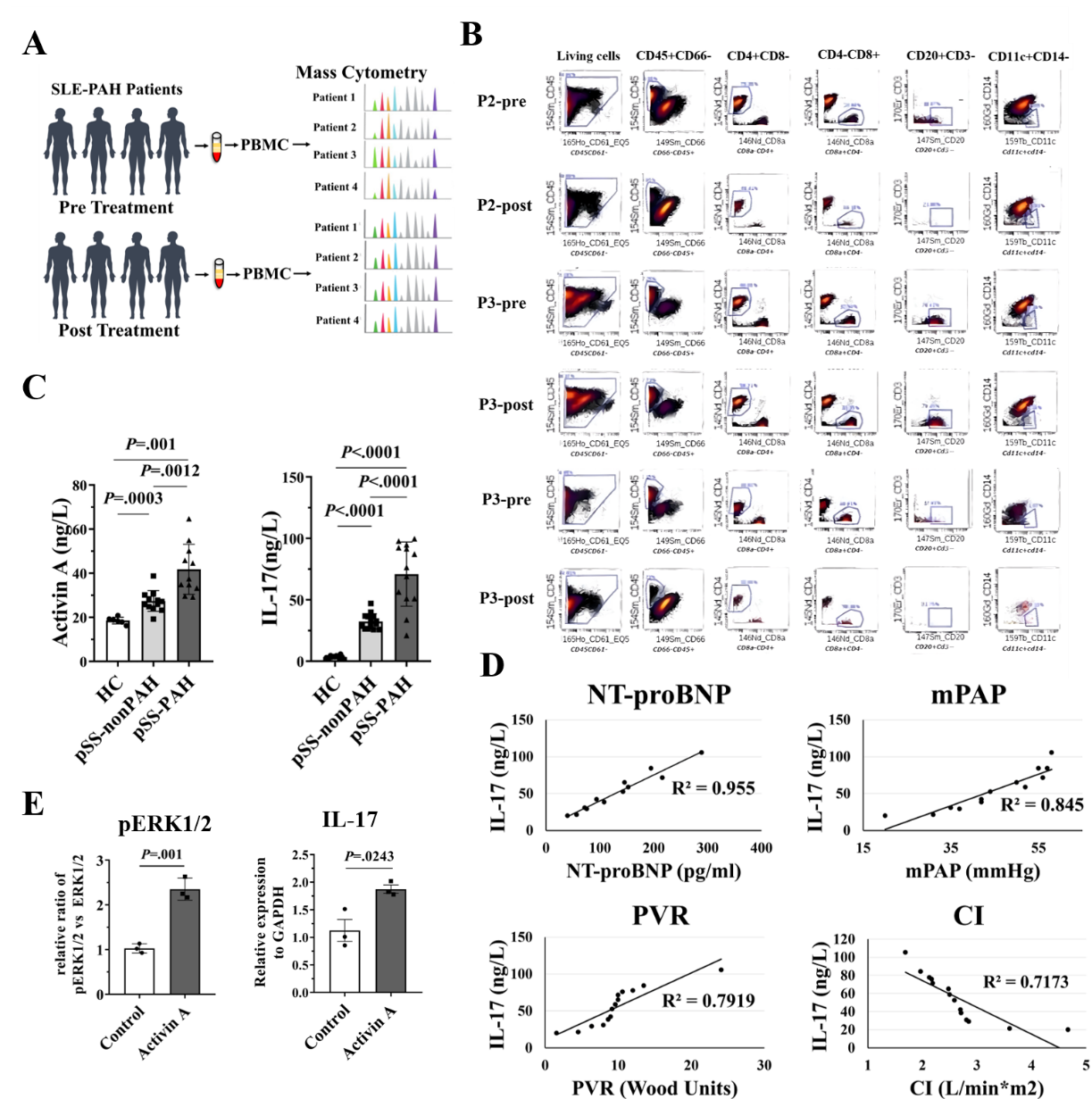

**Figure S1. The percentage of cells of the SLE-PAH patient pre- and post-therapy. (A)** The schematic of Mass Cytometry for four SLE-PAH patients before and after treatment. **(B)** CD4^+^ T cells, CD8^+^ T cells, B cells and Myeloid dendritic cells in the peripheral blood of the SLE-PAH patient pre- and post-therapy analyzed by mass cytometry. **(C)** ELISA Assay were conducted with sera from healthy control (N=19), pSS-nonPAH patients (N=27) and pSS -PAH patients (N=24). **(D)** Correlation analysis between IL-17 level and clinical parameters in patients with SLE-PAH. **(E)** The densitometric quantification of protein levels of pERK1/2 and IL-17 (N=3). Using one-way ANOVA with repeated measures followed by Bonferroni correction for multiple comparisons. Data were shown as mean ±SEM, *P* values (Bonferroni corrected) are depicted in the panels throughout all figures as **P* < .05, ***P* < .01 and ****P* < .001. ns, not significant, *P* > .05. Using one-way ANOVA with repeated measures followed by Bonferroni correction for multiple comparisons. Each data point in the panels represents one independent subject.

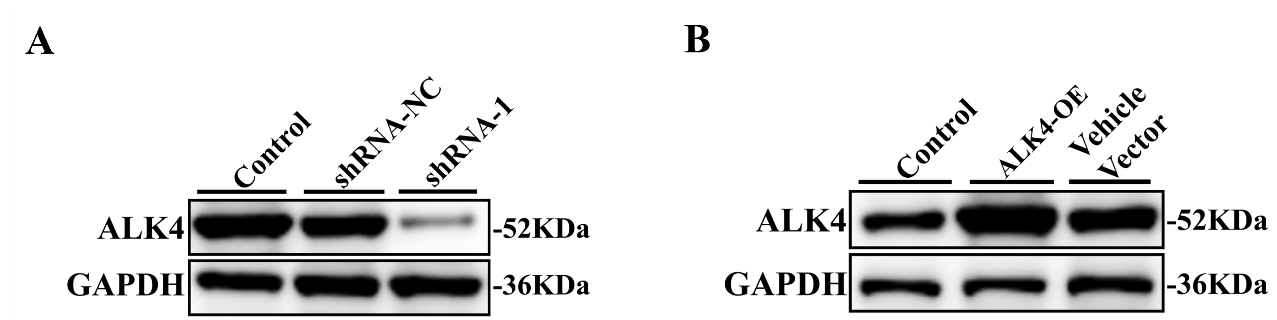

**Figure S2** The effects of ALK4 knockdown by shRNA as well as overexpression in Th17 cells were examined by immunoblotting and the genes expression in lungs of CD4-depleted mice. **(A)** The protein expression of ALK4 in Th17 cells treated with ALK4-shRNA were examined by western blot. **(B)** The protein expression of ALK4 in Th17 cells treated with ALK4-OE lentiviruses were examined by western blot.

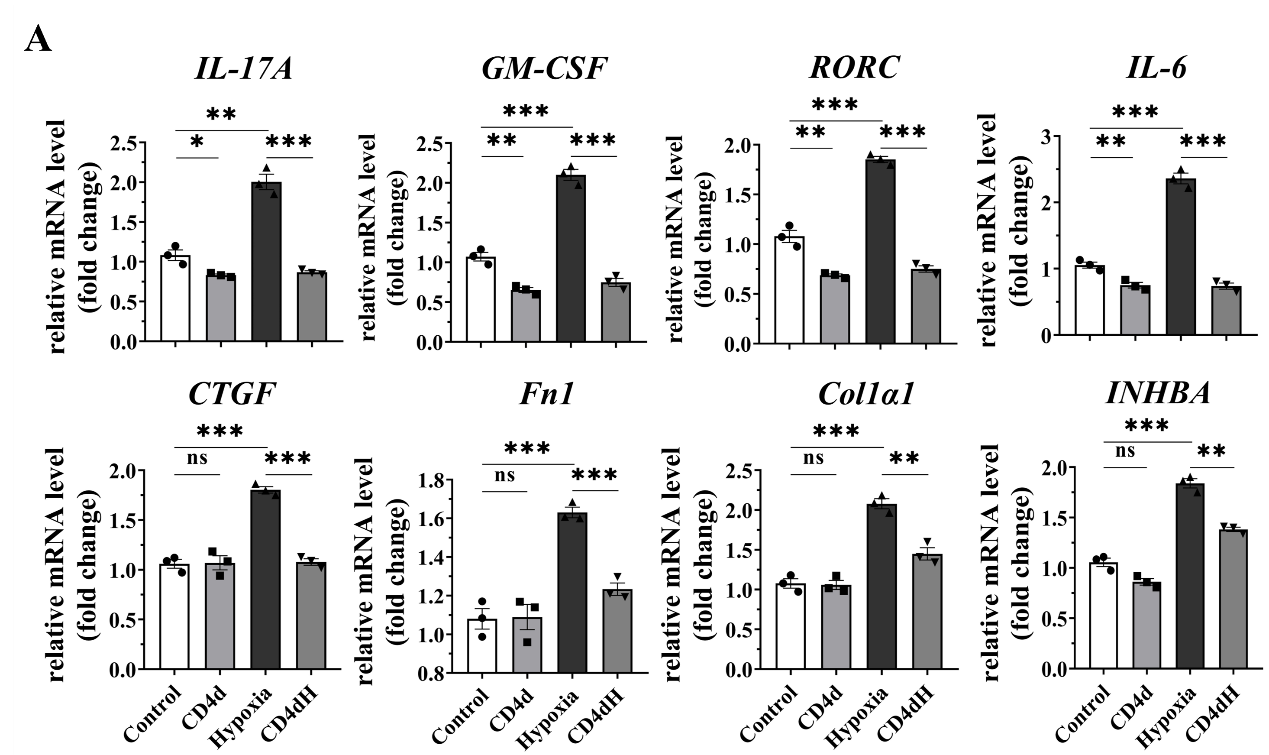

**Figure S3. The genes expression in lungs of CD4-depleted mice. (A)** The genes expression in lungs of CD4-depleted mice. “CD4d” indicated CD4-deleted, “CD4dH” indicated CD4-depleted and hypoxia. Data were shown as mean ±SEM, *P* values (Bonferroni corrected) are depicted in the panels throughout all figures as **P* < .05, ***P* < .01 and ****P* < .001. ns, not significant, *P* > .05. Using one-way ANOVA with repeated measures followed by Bonferroni correction for multiple comparisons. Each data point in the panels represents one independent subject.

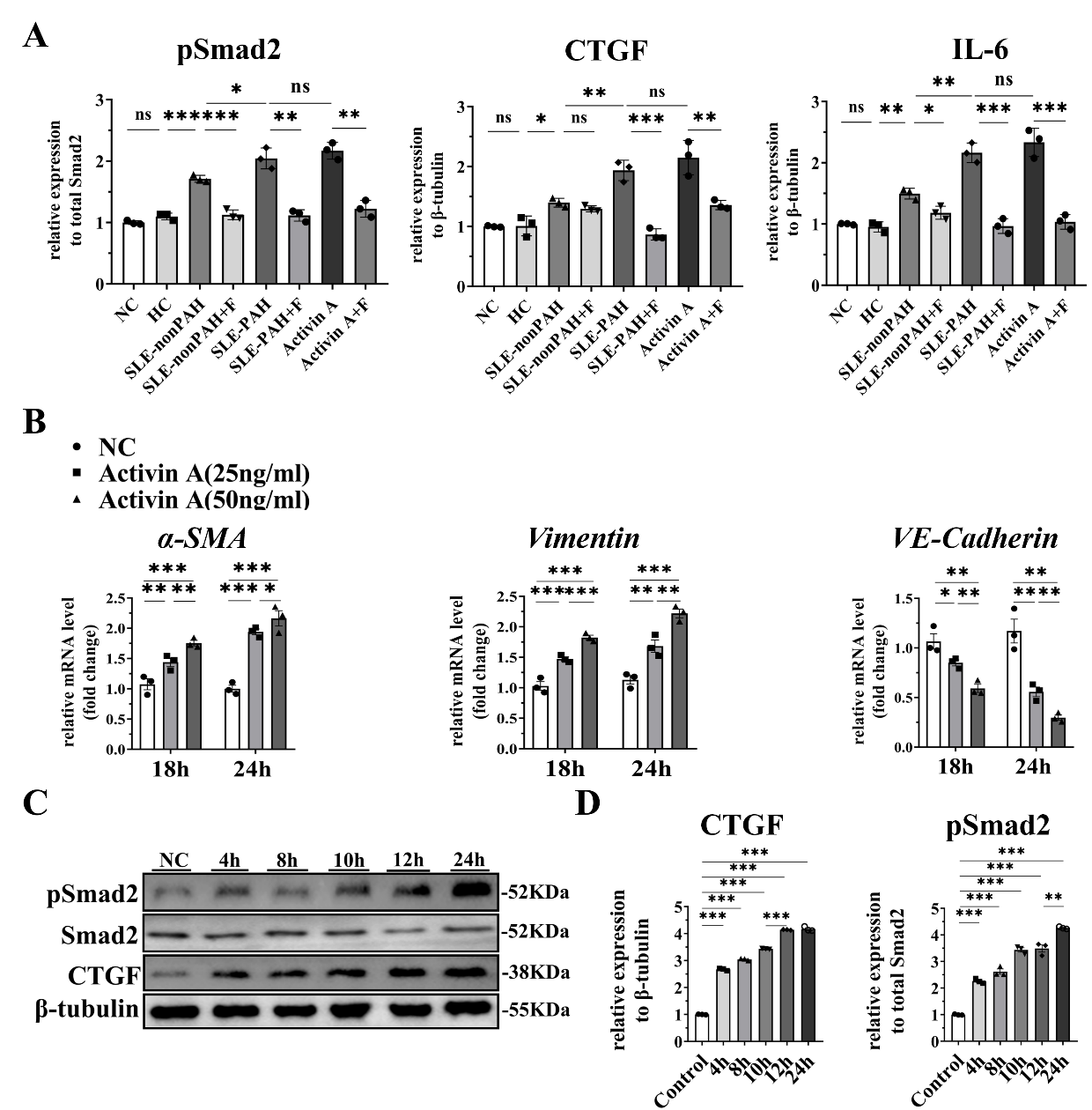

**Figure S4.** The time course of CTGF protein level and Smad2 phosphorylation in hPMECs stimulated with Activin A. **(A)** The densitometric quantification of protein level of pSmad2, CTGF and IL-6 (N=3). **(B)** The mRNA expression of EndoMT markers (*α-SMA*, *Vimentin* and *VE-Cadherin)* were quantified by qPCR in hPMECs induced by Activin A (25ng/ml and 50ng/ml) for indicated time (N=3). **(C)** The protein expression of pSMAD2 and CTGF in hPMECs treated with Activin A(10ng/ml) for indicated time were examined by western blot. **(D)** The densitometric quantification of protein level of CTGF and pSmad2 (N=3). Data were shown as mean ±SEM, *P* values (Bonferroni corrected) are depicted in the panels throughout all figures as **P* < .05, ***P* < .01 and ****P* < .001. ns, not significant. Using one-way ANOVA with repeated measures followed by Bonferroni correction for multiple comparisons. Each data point in the panels represents one independent subject.

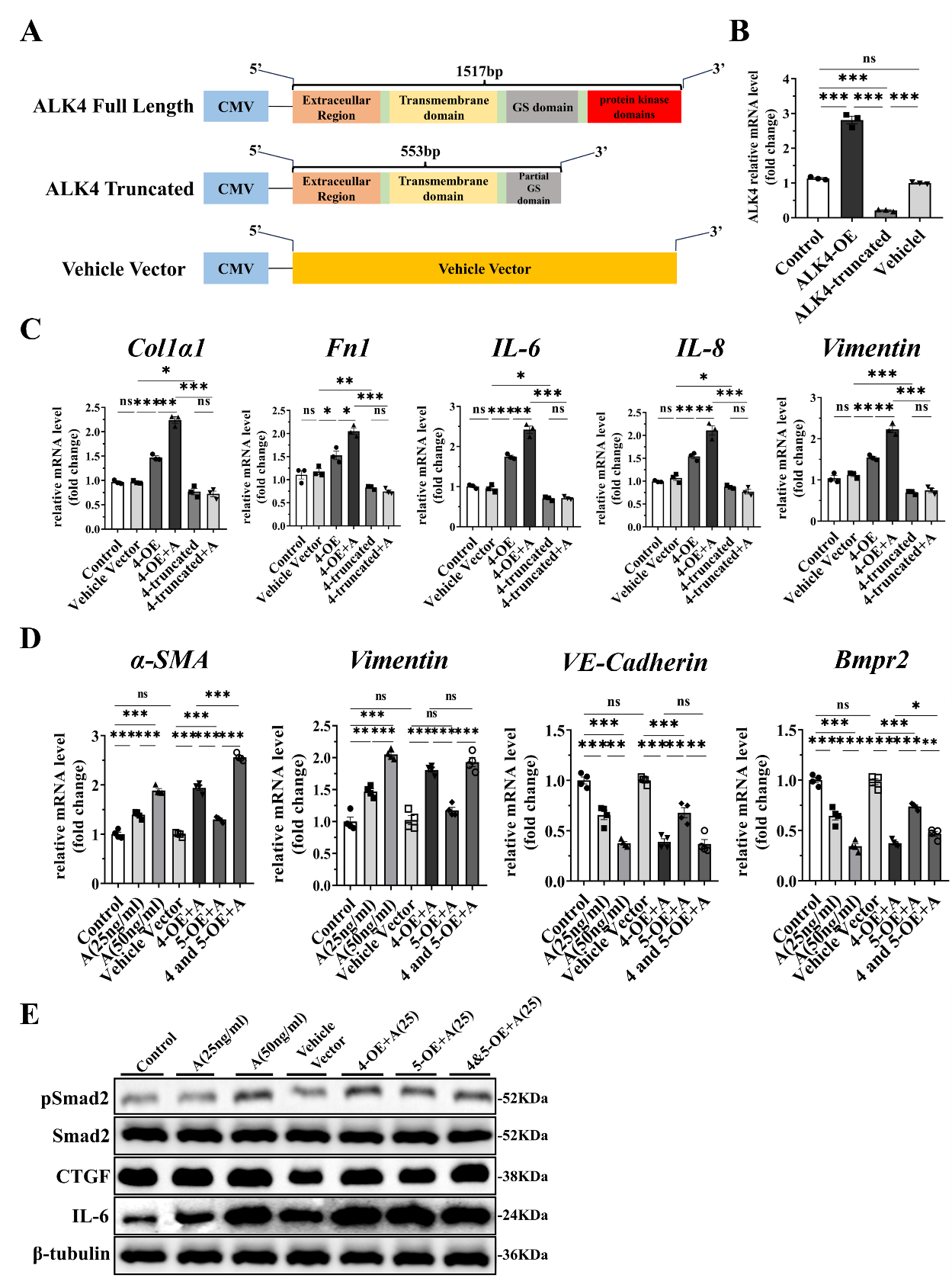

**Figure S5.** The gene expression in hPMECs transfected with ALK4-OE/trancated and ALK5-OE plasmids. **(A)** The schematic diagram showing full length of ALK4, truncated ALK4 and vehicle vector. **(B)** The mRNA expression of ALK4 of hPMECs after treated with ALK4-OE, ALK4- truncated and Vehicle Vector lentiviruses (N=3). **(C)** The mRNA expression of *Col1α1*, *Fn1*, *IL-6*, *IL-8* and *Vimentin* were quantified by qPCR in Control, ALK4-OE, ALK4-truncatedand and Vehicle Vector group (N=3). **(D)** The mRNA expression of *α-SMA*, *Vimentin, VE-Cadherin* and *Bmpr2* in hPMECs transfected with ALK4-OE and ALK5-OE plasmid. OE: overexpression (N=3). **(E)** The protein expression of pSmad2, CTGF and IL-6 in hPMECs transfected with ALK4-OE and ALK5-OE plasmid. Data were shown as mean ±SEM, *P* values (Bonferroni corrected) are depicted in the panels throughout all figures as **P* < .05, ***P* < .01 and ****P* < .001. ns, not significant. Using one-way ANOVA with repeated measures followed by Bonferroni correction for multiple comparisons. Each data point in the panels represents one independent subject.

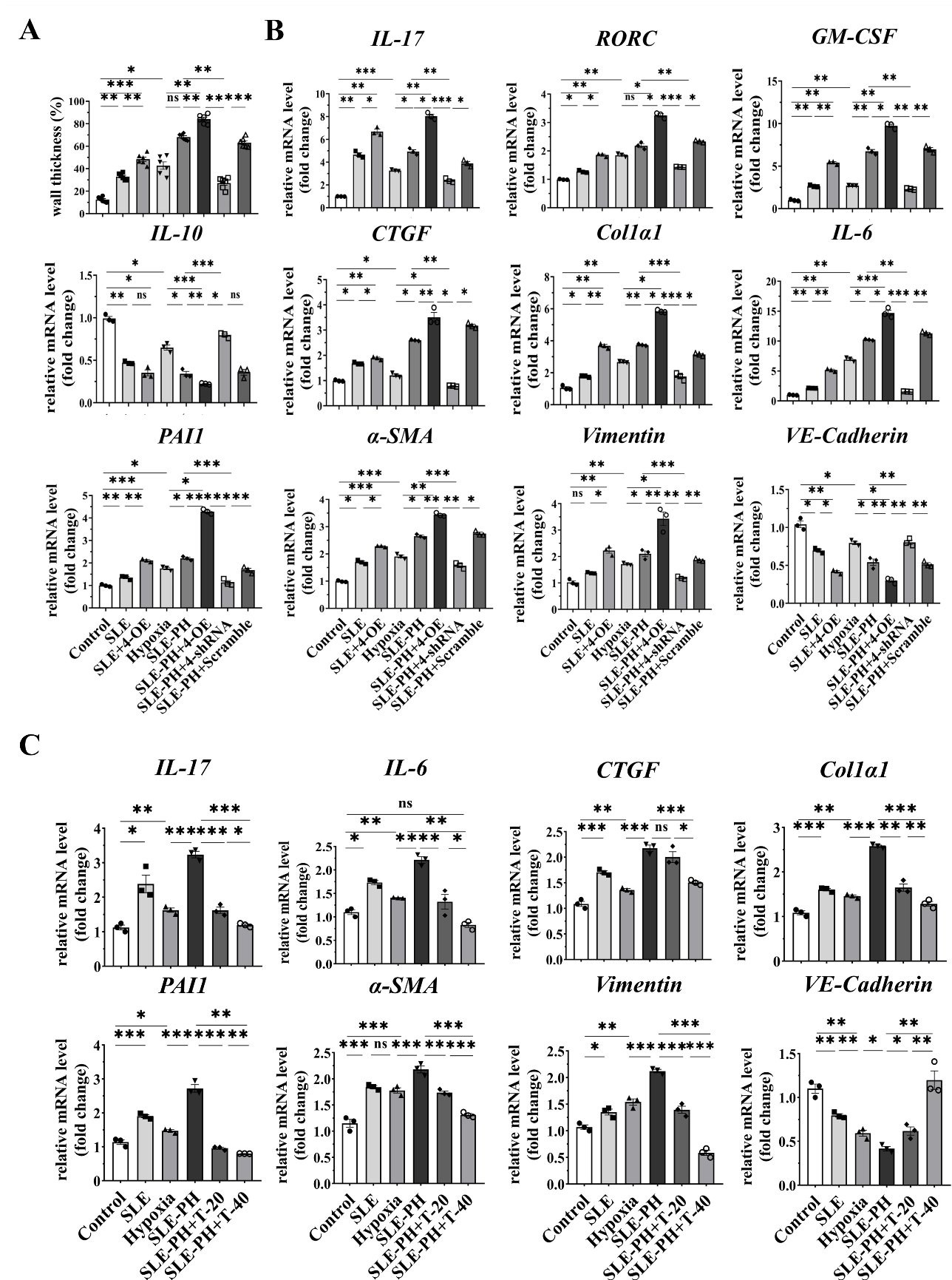

**Figure S6. The genes expression in SLE-PH mouse lungs. (A)** Medial wall thickness of the small pulmonary arteries. **(B)** The mRNA expression in ALK4 overexpression and ALK4 knockdown of SLE-PH mouse model (N=3). **(C)** The mRNA expression in SLE-PH mouse model (N=3). Data were shown as mean ±SEM, *P* values (Bonferroni corrected) are depicted in the panels throughout all figures as **P* < .05, ***P* < .01 and ****P* < .001. ns, not significant. Using one-way ANOVA with repeated measures followed by Bonferroni correction for multiple comparisons. Each data point in the panels represents one independent subject.

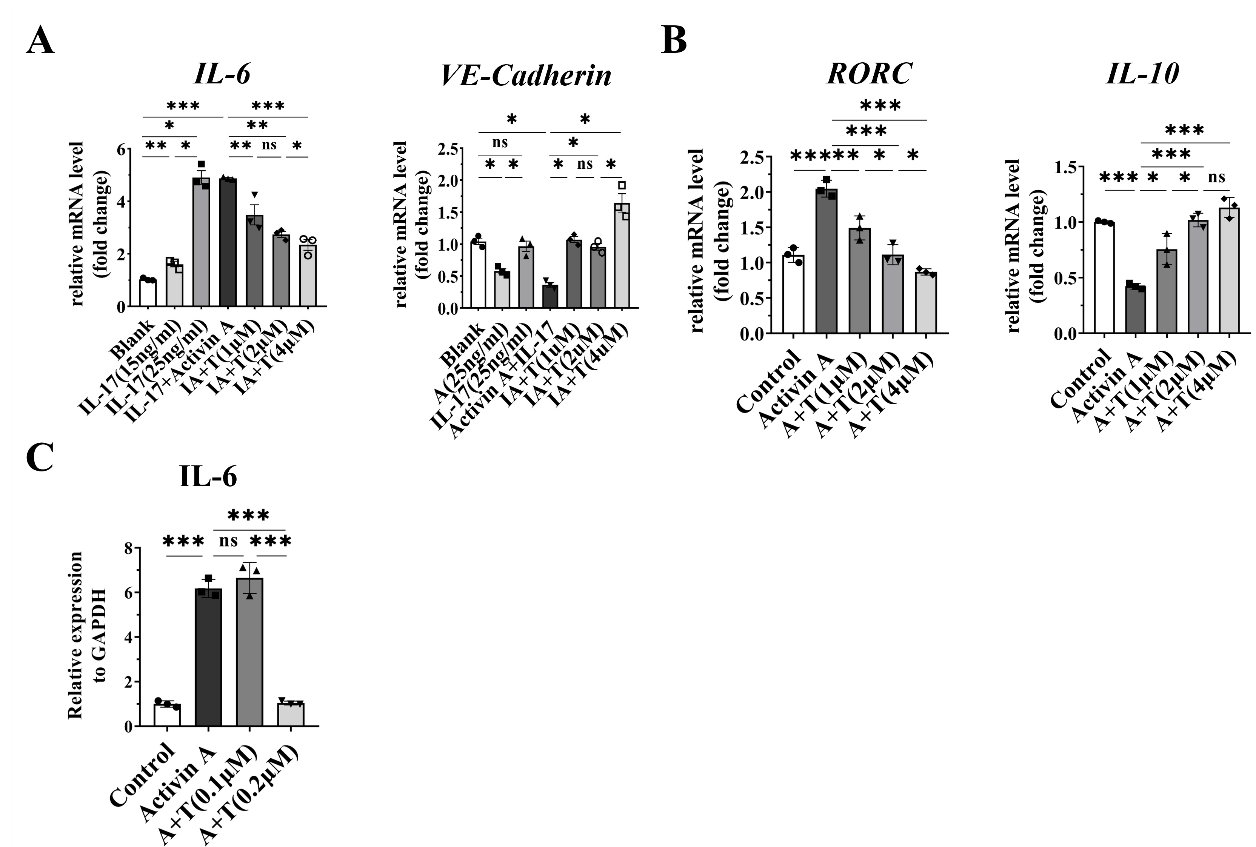

**Figure S7 The effects of TEW on hPMECs and Th17 cells. (A)** The mRNA expression in hPMECs treated with TEW after stimulation with IL-17 and Activin A (N=3). **(B)** The mRNA expression in Th17 cells treated with TEW after stimulation with Activin A (N=3). **(C)** The densitometric quantification of protein levels of IL-6 (N=3). Data were shown as mean ±SEM, *P* values (Bonferroni corrected) are depicted in the panels throughout all figures as **P* < .05, ***P* < .01 and ****P* < .001. ns, not significant. Using one-way ANOVA with repeated measures followed by Bonferroni correction for multiple comparisons.

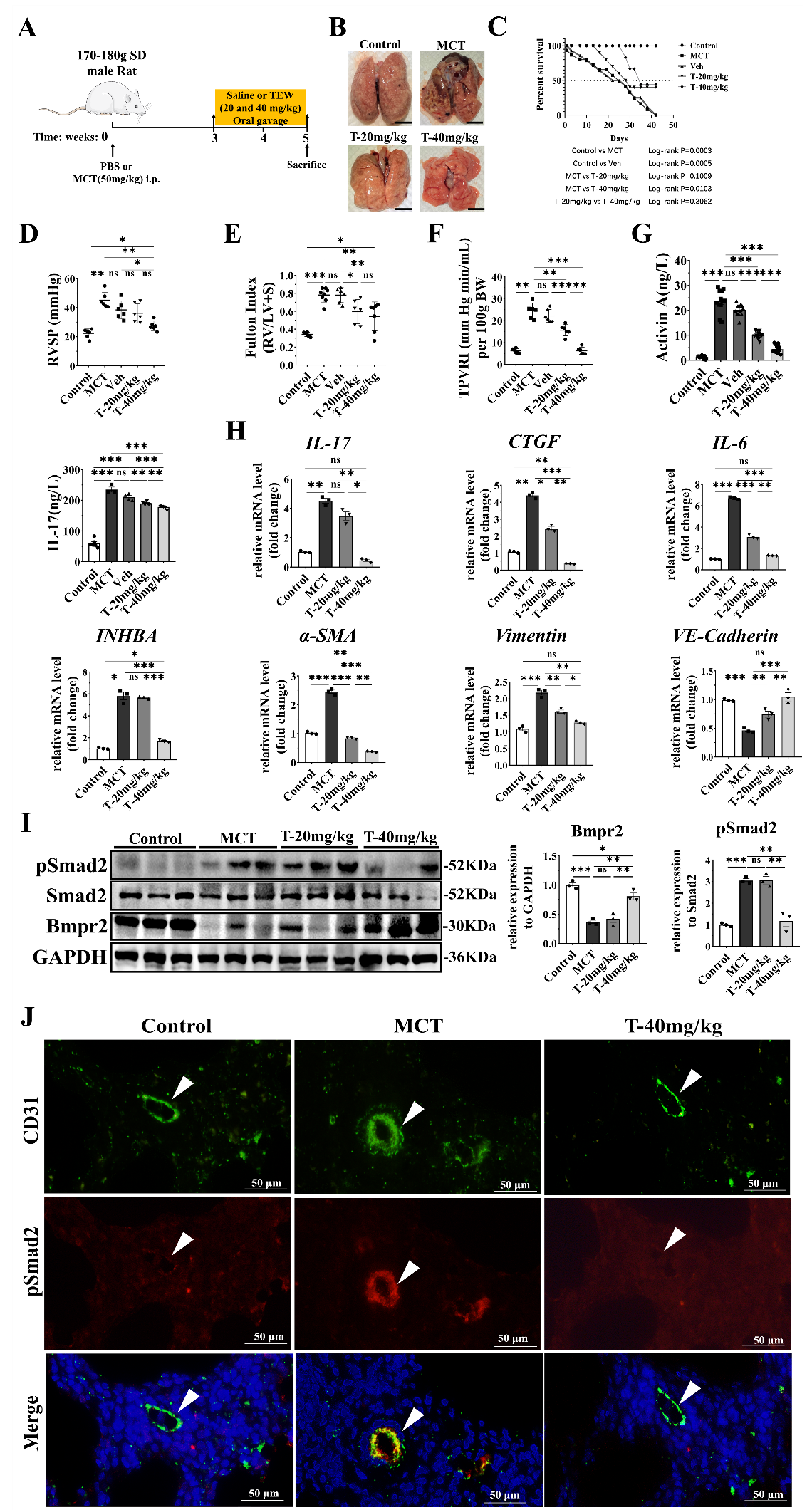

**Figure S8. Therapeutic effect of TEW in MCT-induced Rat PAH Model. (A)** The Schematic Illustration rat pulmonary hypertension model generated by MCT. **(B)** Representative photograph of the whole lung (Scale bars: 1cm). **(C)** The survival rates of MCT-induced Rat PAH Model. **(D)** RVSP. **(E)** Fulton index. **(F)** TPVRI. (N=6 rats for each group). **(G)** Serum level of Activin A and IL-17 in MCT-induced Rat PAH Model. **(H)** The mRNA expression of lungs in MCT-induced Rat PAH Model (N=3). **(I)** The protein expressions of Bmpr2 and pSmad2 in lungs from each group were examined by western blot and densitometric quantification was shown in right (N=3). “T-40” indicated TEW (40mg/kg). The samples derive from the same experiment and that blots were processed in parallel. **(J)** Representative images showed CD31 (green) and pSmad2 (red) with DAPI (blue) in MCT-induced rat PAH model assessed by immunofluorescence microscopy from the indicated groups. White arrow indicates the pulmonary small arterial. Scale bar=50 μm. Data were shown as mean ±SEM, *P* values (Bonferroni corrected) are depicted in the panels throughout all figures as **P* < .05, ***P* < .01 and ****P* < .001. ns, not significant. Using one-way ANOVA with repeated measures followed by Bonferroni correction for multiple comparisons. Each data point in the panels represents one independent subject.

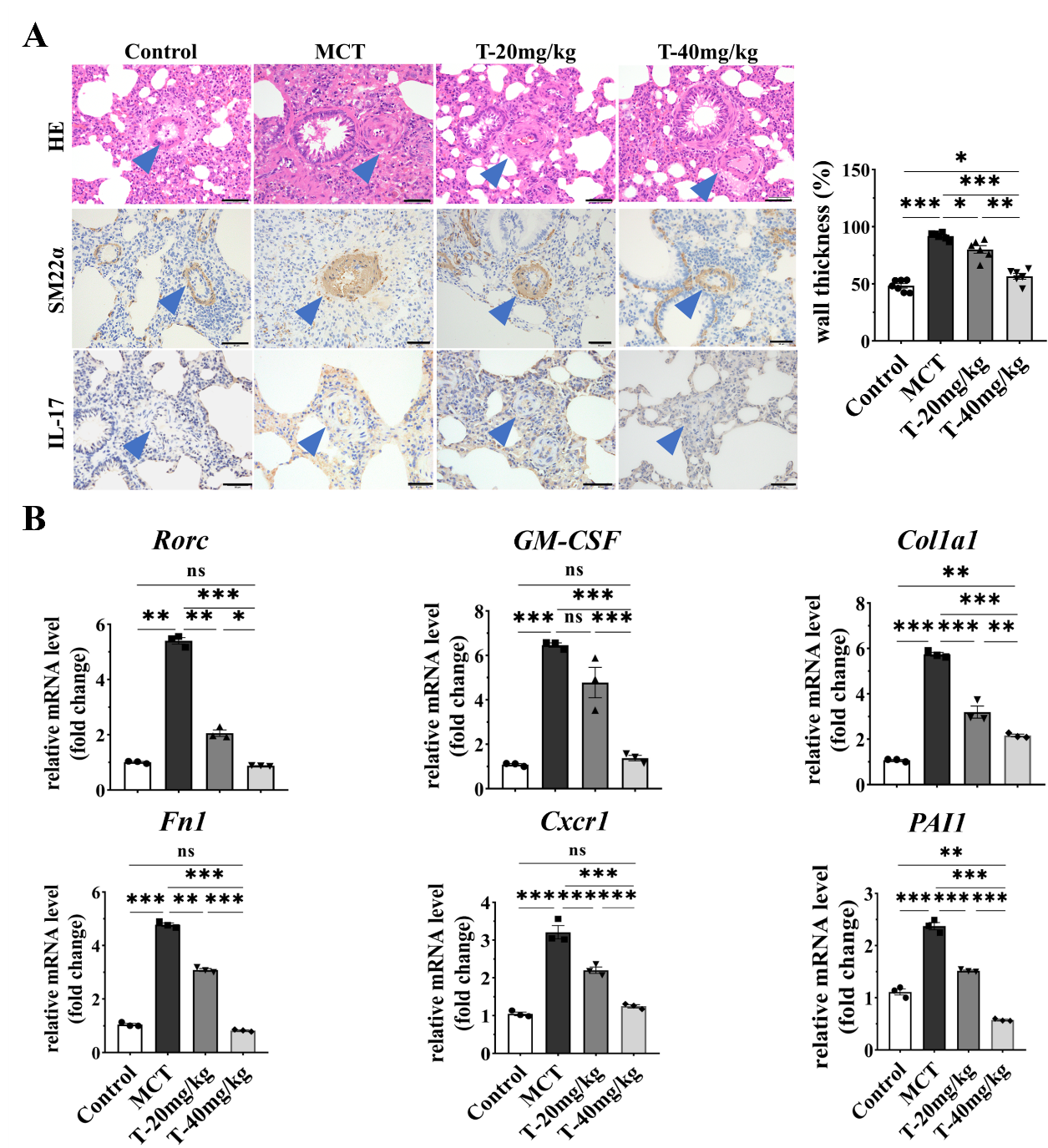

**Figure S9.** **The reversed pulmonary vascular remodeling and genes expression by TEW in MCT-induced Rat PAH model.** **(A)** The representative images of pulmonary arteries (Scale bar=50 μm) by HE staining and IL-17 IHC staining of each group indicated in the panel and medial wall thickness of the small pulmonary arteries (N=6). **(B)** The mRNA expression in MCT-induced Rat PAH Model (N=3). Data were shown as mean ±SEM, *P* values (Bonferroni corrected) are depicted in the panels throughout all figures as **P* < .05, ***P* < .01 and ****P* < .001. ns, not significant. Using one-way ANOVA with repeated measures followed by Bonferroni correction for multiple comparisons. Each data point in the panels represents one independent subject.

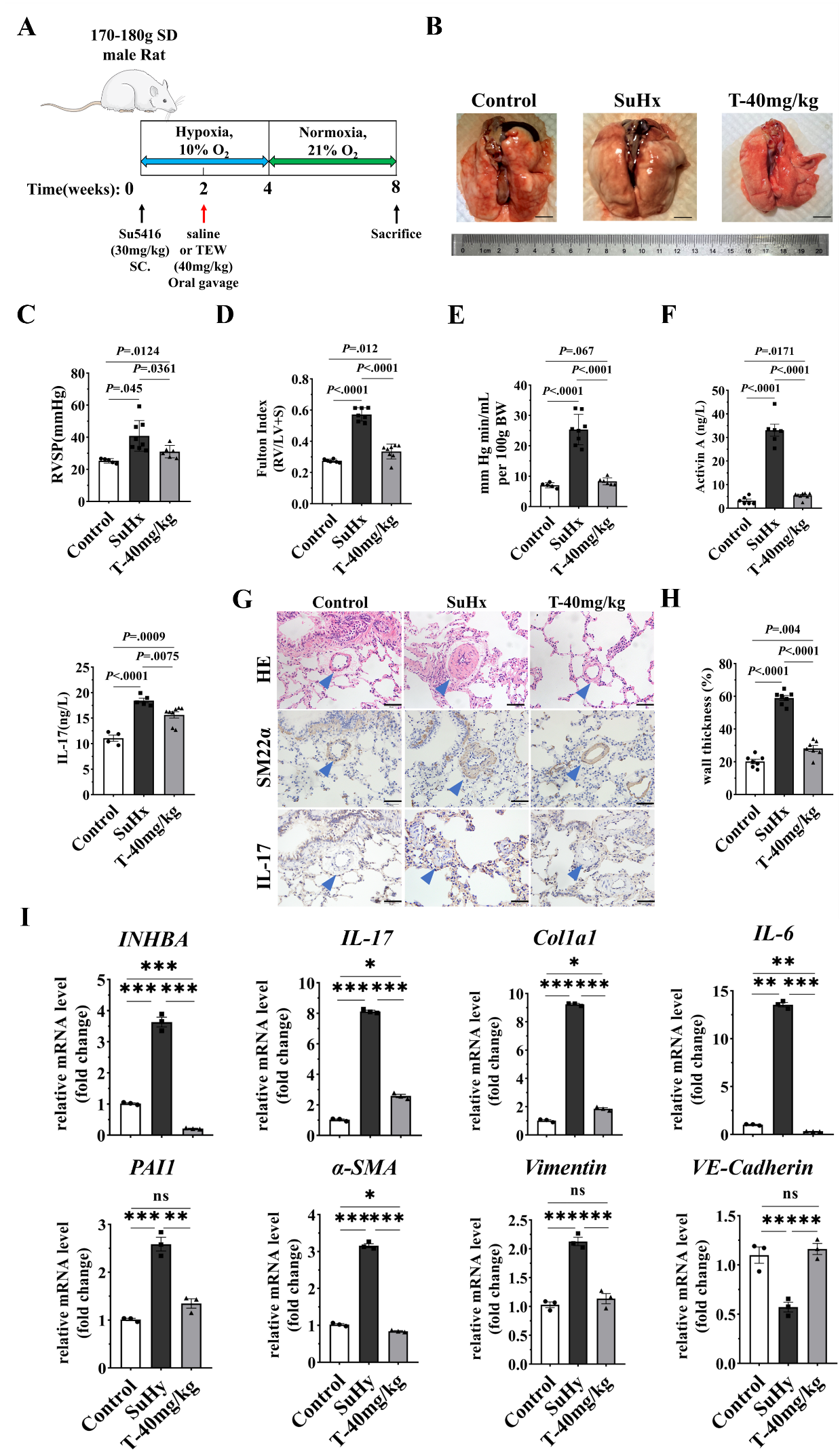

**Figure S10.** **TEW Treatment Ameliorated SuHx PAH in Rat.** **(A)** The workflow of sugen5416- induced hypoxia mouse pulmonary hypertension model. **(B)** Representative images of the whole lung (Scale bars: 1cm). **(C)** RVSP. **(D)** Fulton index. **(E)** TPVRI. (N=6-8 rats for each group). **(F)** Serum level of IL-17 and Activin A in SuHx PAH Rats. **(G)** The representative images of pulmonary arteries (Scale bar=50 μm) by HE staining, SM22α and IL-17 of each group indicated in the panel. Blue arrow indicates the pulmonary small arterial. **(H)** Medial wall thickness of small pulmonary arteries. **(I)** The mRNA expression in SuHx Rat PAH Model (N=3). Data were shown as mean ±SEM, *P* values (Bonferroni corrected) are depicted in the panels throughout all figures as **P* < .05, ***P* < .01 and ****P* < .001. ns, not significant. Using one-way ANOVA with repeated measures followed by Bonferroni correction for multiple comparisons. Each data point in the panels

represents one independent subject.

**Supplemental Table 1**

| Patients | Sex | Age, years | WHO  Classification | Treatment | Outcome |
| --- | --- | --- | --- | --- | --- |
| SLE-PAH-1 | female | 30 | II | Prednisone 1mg/kg/d, tapered to 7.5mg/d maintained, Tacrolimus 1mg bid, Hydroxychloroquine  0.2g bid, Ambrisentan 5mg qd | Responder,  PAH low-risk reached |
| SLE-PAH-2 | female | 25 | II | NA | NA |
| SLE-PAH-3 | female | 52 | II | Prednisone 1mg/kg/d, tapered to 5mg/d maintained, Cyclophosphamide  0.4g qw for six months, Hydroxychloroquine  0.2g bid, Bosentan  125mg bid | Responder,  PAH low-risk reached |
| SLE-PAH-4 | female | 30 | II | Prednisone 1mg/kg/d, tapered to 5mg/d maintained, Cyclophosphamide  0.4g qw for six months, Hydroxychloroquine  0.2g bid | Responder,  PAH low-risk reached |

The treatments and outcome of SLE-PAH patients for mass cytometry

**Supplemental Table 2**

Clinical Characteristics of representative patients with SLE-PAH

| Variable | SLE-nonPAH Patients  (N=9) | SLE-PAH Patients  (N=14) |
| --- | --- | --- |
| Sex, no. female/male | 7/2 | 14/0 |
| Age, years | 39.0±9.6 | 41.4±9.4 |
| WHO Classification |  |  |
| Class II | NA | 14 |
| Class III | NA | 0 |
| 6 min walk distance, m | NA | 518.1±52.7 |
| NT-proBNP, pg/ml | NA | 118.2±57.0 |
| mPAP, mm Hg | NA | 45.7±12.0 |
| PVR, Wood Units | NA | 9.8±5.1 |
| CI, L/min*m^2^ | NA | 2.6±0.7 |

*The mPAP and PVR were determined by right side heart catheterization when PAH was diagnosed. NT-proBNP = N-Terminal Pro-Brain Natriuretic Peptide; mPAP = mean pulmonary arterial pressure; PVR = pulmonary vascular resistance; CI = cardiac index. NA = not available. The total number of healthy controls is 20, including 18 females and 2 males. The mean age of healthy controls is 27.7 (SD, 5.2) years old.

### Reference

1.	Attisano L, Wrana JL, Montalvo E, Massague J. Activation of signalling by the activin receptor complex. Mol Cell Biol 1996;16:1066-73.

2.	Xing Y, Zhao S, Wei Q, Gong S, Zhao X, Zhou F, et al. A novel piperidine identified by stem cell-based screening attenuates pulmonary arterial hypertension by regulating BMP2 and PTGS2 levels. Eur Respir J 2018;51.
